# The potential impact of vaccine passports on inclination to accept COVID-19 vaccinations in the United Kingdom: evidence from a large cross-sectional survey and modelling study

**DOI:** 10.1101/2021.05.31.21258122

**Authors:** Alexandre de Figueiredo, Heidi J. Larson, Stephen Reicher

## Abstract

**Background:** Four vaccines against the novel coronavirus 2019 disease (COVID-19) caused by the severe acute respiratory coronavirus 2 (SARS-CoV-2) have currently been approved for use in the United Kingdom. As of 30 April 2021, over 34 million adults have received at least one dose of a COVID-19 vaccine. The UK Government is considering the introduction of vaccine passports for domestic use and to facilitate international travel for UK residents. Although vaccine incentivisation has been cited as a motivating factor for vaccine passports, it is currently unclear whether vaccine passports are likely to increase inclination to accept a COVID-19 vaccine.

**Methods:** We conducted a large-scale national survey in the UK of 17,611 adults between 9 and 27 April 2021. Bayesian multilevel regression and poststratification is used to provide unbiased national-level estimates of the impact of the introduction of vaccine passports on inclination to accept COVID-19 vaccines among all respondents who have not yet had two vaccination doses. Multilevel regressions identify the differential impact of the likely impact of vaccine passports on uptake intent between socio-demographic groups. Gibbs sampling was used for Bayesian model inference, with 95% highest posterior density intervals used to capture uncertainty in all parameter estimates.

**Findings:** We find that the introduction of vaccine passports will likely lower inclination to accept a COVID-19 vaccine once baseline vaccination intent has been adjusted for. Notably, this decrease is larger if passports were required for domestic use rather than for facilitating international travel. The impact of passports while controlling for baseline vaccination intent differentially impacts individuals by socio-demographic status, with being male (OR 0·87, 0·76 to 0·99) and having degree qualifications (OR 0·84, 0·72 to 0·94) associated with a decreased inclination to vaccinate if passports were required for domestic use, while Christians (OR 1·23, 1·08 to 1·41) have an increased inclination over atheists or agnostics. There is a strong association between change in vaccination inclination if passports were introduced and baseline vaccination intent: stated change in vaccination inclination is thus lower among Black or Black British respondents (compared to Whites), younger age groups, and non-English speakers. We find notable sub-national trends, for example, that passports could increase inclination among students and Jewish respondents in London compared to those in full-time education or atheists or agnostics, respectively.

**Interpretation:** To our knowledge, this is the first quantitative assessment of the potential impact of the introduction of vaccine passports on COVID-19 vaccine intention. Our findings should be interpreted in light of sub-national trends in current uptake rates across the UK, as our results suggest that vaccine passports may induce a lower vaccination inclination in socio-demographic groups that cluster geographically in large urban areas. Caution should therefore be exercised in introducing passports as they may result in less positive health-seeking behaviours for the COVID-19 vaccine (as well as other existing or future vaccinations) and may contribute to concentrated areas of low vaccinate uptake, which is an epidemic risk. We call for further evidence on the impact of vaccine certification on confidence in COVID-19 vaccines and in routine immunisations in wider global settings and, in particular, in countries with low overall trust in vaccinations or in authorities that administer or recommend vaccines.

**Funding:** This survey was funded by the Merck Investigator Studies Program (MISP)

**Research in context:** *Evidence before this study:* Proof of vaccination has, to date, had limited use in public and private settings for the UK public, such as proof of yellow fever vaccination for international travel to limited destinations, or requirements of Hepatitis B vaccination in some medical roles. Although recent surveys have suggested that the majority of the British public support vaccine passports, we are not aware of any studies assessing the impact that proof of vaccination status for domestic use or for international travel may have on vaccination inclination and thus—perhaps more importantly—on epidemic spread.

*Added value of this study:* We conducted a large-scale survey of more than 17,000 members of the UK public to explore attitudes to vaccine passports for domestic and international use. Bayesian methods are used to compute nationally representative estimates of the impact of vaccine passports on change in inclination to accept COVID-19 vaccines and to establish the socio-demographic determinants of vaccination inclination. This study is, as far as we are aware, the first to assess the impact of vaccine passports on vaccination inclination in the UK.

*Implications of all the possible evidence:* This study provides novel insights into the potential impact of vaccine passports on COVID-19 vaccine intent in the UK. Although we find that vaccine passports receive popular support in the UK, there exists large variations in their appeal that stratify along socio-demographic lines. Most notably, younger age groups, Black and Black British ethnicities (compared to Whites), and non-English speakers are more likely to express a lower inclination to vaccinate if passports were introduced. Although these groups comprise a relatively small proportion of the UK population, there are crucial issues that these perceptions among these groups cause: notably, that these groups tend to have lower baseline vaccination intent and they cluster geographically. Therefore, since geographic clusters of low vaccination uptake can result in disproportionate increases in required vaccination levels for herd immunity in adjacent settings, we need to exercise extreme caution in public health interventions that may push these areas further away from vaccination. This is especially so if such an intervention will have little overall impact on the majority of the population outside these areas who are already quite willing to vaccinate. Overall, we find that the introduction of passports for either domestic or international use has a net negative impact on vaccination inclination, once we control for baseline vaccination intent. Our findings suggest that vaccination passports may not only yield damaging health outcomes for already marginalised communities: this may lead to further distrust in the government and public health systems and may have negative downstream consequences for other health-seeking behaviours, for example, routine immunisations.

## Introduction

Proof of vaccination status via an electronic or physical vaccine passport or certificate has been proposed as a means to aid in the reopening of society after the implementation of non-pharmaceutical intervention^1–3^. The discussion around the use of vaccine passports for domestic use in the United Kingdom (UK) has largely centred on their use in non-medical social settings where physical distancing may be challenging, such as public houses, restaurants, and large sporting events. Vaccine passports have also been proposed as a means to speed up the reopening of international travel for freedom of movement or tourism^2^, which has largely halted over the past year due to various restrictions on international travel.

There has been much debate about the relative merits of vaccine passports, with incentivising vaccination^4^, public health principles of least infringement^5^ (though arguments have also been made that passports could be more restrictive^6^), and minimising SARS-CoV-2 risk when reopening society^3^ cited as arguments in favour of vaccine passports. Major ethical concerns remain, however^5^. It has been argued that requiring proof of vaccination to re-enter society may violate freedom of choice^7^ and further suggested that a requirement to vaccinate to fully re-enter society may stigmatise those who opt not to vaccinate^5,7^ (who may stratify along socio-demographic characteristics leading, ultimately, to barriers or unequal treatment between socio-demographic groups^5,8,9^) or penalising those who opt not to receive vaccination healthcare through financial and logistical costs to prove disease or immunological status via SARS-CoV-2 tests or antibody tests (the results of will likely be recorded on any vaccine passport or certificate that is introduced). As vaccinated individuals may still be infected with COVID-19—and the current level of sterilising immunity from COVID-19 vaccines is unclear^10^—vaccine passports may lead to excluding healthy non-infected or immune individuals from societal events while infectious vaccinated individuals fully return.

These ethical concerns and potential additional costs must be considered in a contextual basis for policy in the UK, where confidence in, and uptake of, routine immunisations strongly depend on socio-demographic status^11–13^. With regards to COVID-19 vaccines in particular, females, younger age groups, Black / Black British ethnicities, Muslims, and Polish speakers have been less likely to state intent to vaccinate compared to males, older age groups, Whites, atheists or agnostics, and English or Welsh speakers (respectively)^14^. Early evidence from observed uptake in the UK suggests that gender and ethnicity are associated with lower uptake among healthcare workers^15^, and non-Whites have lower uptake than Whites among the general population too, with Black African and Black Caribbean people with the lowest uptake across the UK^16,17^. Although specific reasons for hesitancy will vary both within and between these groups, trust in authorities and the Government, as well as historic marginalisation^18^, play a key role^19^ and it is currently unclear how the introduction of health status passports or certificates will affect intent to vaccinate as well as a breakdown of trust in authorities recommending vaccinations.

Recent polling in the UK has suggested that vaccine passports receive majority support in the UK, with increased support for use in international travel^20,21^. In this study, we quantitively assess the likely impact of the introduction of vaccine passports for domestic and international use on inclination to accept COVID-19 vaccines using a large nationally representative cross-sectional survey of about 17,000 UK adults conducted in April 2021. In particular, we seek to establish whether vaccine passports are likely to encourage or discourage uptake of COVID-19 vaccines among people who have not yet had two doses of a COVID-19 vaccine. We compute the overall impact of the introduction of vaccine passports on intent to vaccinate and identify the differential impact of passports on vaccination intent across socio-demographic and across UK region.

## Methods

### Data collection and processing

A total of 17,611 adults were surveyed between 9 April 2021 and 27 April 2021. Respondent quotas were set to match UK national demographic counts by sex, age, and sub-national region. During data collection, quality control procedures resulted in the removal of 1,084 responses (see appendix). All respondents were recruited via online panels by ORB (Gallup) International (www.orb-international.com). Informed consent was obtained by all respondents before respondents participated in the survey (see appendix for the full survey questionnaire which includes the informed consent statement presented to participants).

Respondents were asked ‘*If a coronavirus (COVID-19) certificate or passport was required* ***to attend social events in the UK*** *(such as sports events, theatres, pubs, or restaurants), would you be more or less inclined to accept a coronavirus (COVID-19) vaccine?’* and ‘*If a coronavirus (COVID-19) certificate or passport was required* ***for international travel***, *would you be more or less inclined to accept a coronavirus (COVID-19) vaccine?*’ (emphasis added). As many respondents may not be aware of vaccine passports or certificates, a brief definition was first given (see appendix). Responses are given on a five-point scale: ‘*much less likely*’, ‘*somewhat less likely*’, ‘*neither more nor less likely*’, ‘*somewhat more likely*’, or ‘*much more likely*’ and assigned a numeric value from 1 to 5, respectively. Before being asked to report change in vaccine inclination, respondents were primed with a brief definition of vaccine passports: ‘*We would now like to ask you some questions about a vaccine or immunity certificate (commonly referred to as a “vaccine passport”). A vaccine or immunity certificate is a physical or electronic document that confirms your status against a particular disease. For example, the certificate could confirm that you have been vaccinated against a disease or that you have some pre-existing immunity’*.

In exploring change in vaccination intent if vaccine passports were required for domestic or international use, it is important to control for individuals’ baseline level of vaccination intent, as survey questions that investigate how an information or event changes their attitudes may illicit the same response as the underlying attitude being measured itself^22^. Thus, to control for existing vaccination intent, respondents are first asked whether they have been offered a COVID-19 vaccine and, if so, whether they have taken the vaccine (and how many doses). Respondents who reported taking one dose only were asked whether they intended on receiving a second dose (‘*Do you intend on receiving your second dose?*’), while respondents who reported not having received the vaccine were asked if they intended on accepting the vaccine (*‘Do you intend on accepting a coronavirus (COVID-19) vaccine?’*). All respondents who have not been invited to vaccinate were asked whether they would take the vaccine (‘*When you are invited to take a coronavirus (COVID-19) vaccine, will you accept the vaccine for yourself?*’) (figure 1). Responses to all preceding questions in parentheses could answer on the four-point scale, ‘*yes, definitely’*, ‘*unsure, but leaning towards yes*’, ‘*unsure, but leaning towards no*’, or ‘*no, definitely not*’ and were collated into a single variable. These responses are assigned the values 1 to 4, respectively. As we wish to explore the impact of vaccine passports on future vaccination inclination, all respondents who have already received both doses (1,984, figure 1) were removed from entirely from all analyses.

**Figure 1.**
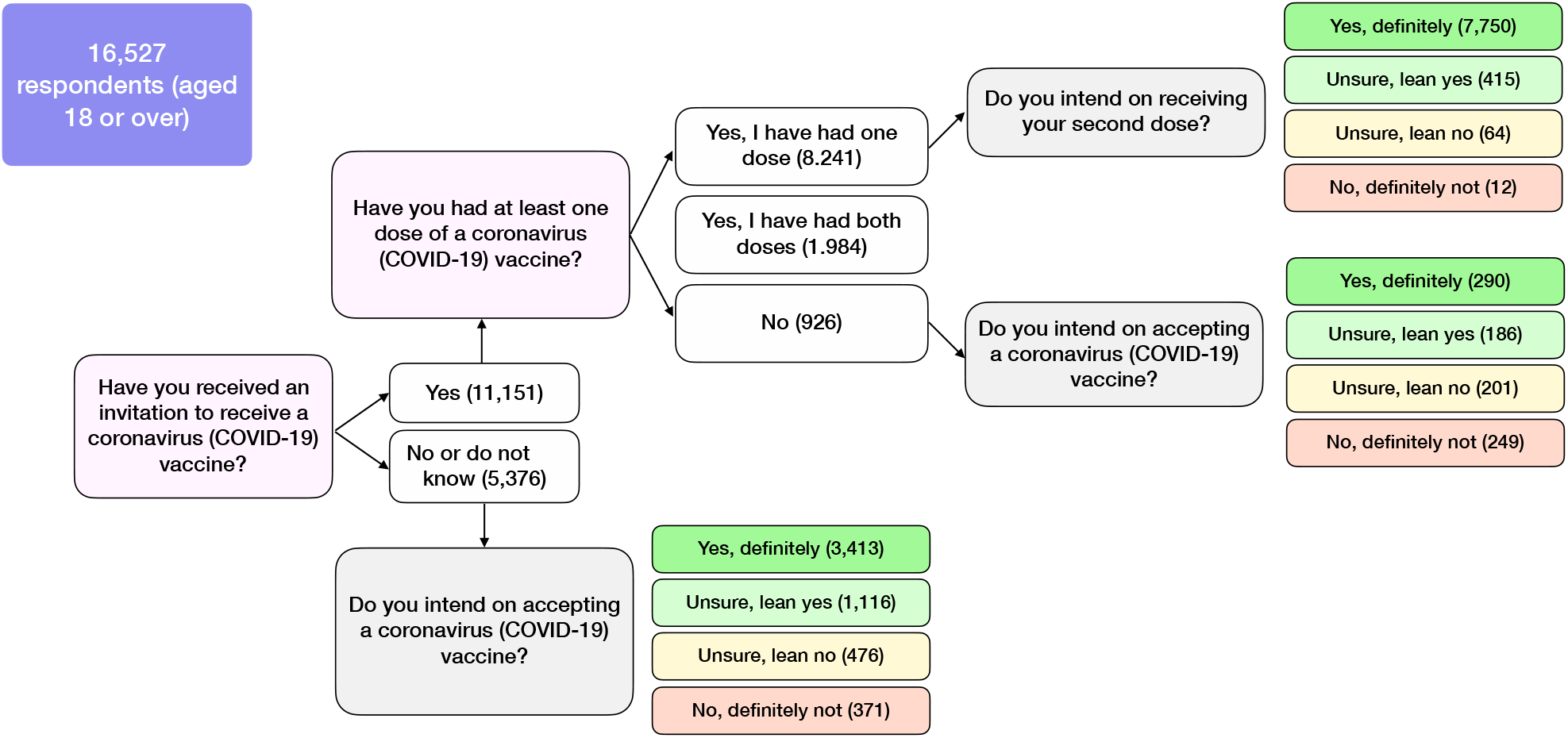
Baseline intent to accept a COVID-19 vaccine, *Z*.

Individuals’ outer postcode, sex, age, highest level of education, employment status, religious affiliation, ethnicity, and primary language are also recorded. Outer postcode (the first half of a UK postcode) was re-coded to administrative region (the NUTS1 unit, see https://www.ons.gov.uk/methodology/geography/ukgeographies/eurostat). These socio-demographic variables serve two purposes: i) they allow a meaningful exploration of the factors associated with vaccination intent and ii) they align with socio-demographic data collected in the latest census. This latter point allows individual-level re-weighting according to millions of UK census records. All summary of socio-demographic variables used and details on variable recoding are provided in table 1. A breakdown of individuals’ change in vaccination inclination by baseline vaccination intent and socio-demographic group is provided in appendix, table 1 for all individuals who have not had two COVID-19 doses.

**Table 1.** Study data. Survey items are shown with possible responses (including recodes, if any), and baselines used in the ordinal logistic regressions.

|  | Survey question | Values (recodes in parenthesis) | baseline |
| --- | --- | --- | --- |
| Change in COVID-19 vaccination intent | If a coronavirus (COVID-19) certificate or passport was required to attend social events in the UK (such as sports events, theatres, pubs, or restaurants), would you be more or less inclined to accept a coronavirus (COVID-19) vaccine? (Y) | much less inclined (1); somewhat less inclined (2), neither more nor less inclined (3); somewhat more inclined (4); much more inclined (5); do not know / prefer not to say (3) | n/a [ordinal response variable] |
|  | If a coronavirus (COVID-19) certificate or passport was required for international travel, would you be more or less inclined to accept a coronavirus (COVID-19) vaccine? (Y) |  |  |
| COVID-19 vaccination intent | Do you intend on receiving your second dose? [if respondent has had the first dose] OR Do you intend on accepting a coronavirus (COVID-19) vaccine? [if respondent has been invited but not reported at least one dose] OR When you are invited to take a coronavirus (COVID-19) vaccine, will you accept the vaccine for yourself? [If respondent not yet invited to vaccinate] (Z) | Yes, definitely (4); Unsure, but leaning towards yes (3); Unsure, but leaning towards no (2); No, definitely not (1) | n/a [ordinal covariate] |
| Covariates | sex | male and female | female |
|  | age | integer value mapped to 18-24, 25-34, 35-44, 45-54, 55-64, 65-79, 80+ | 18-24 |
|  | highest educational attainment | No academic qualifications (none/other)<br>0-4 GCSE, O-levels, or equivalents (level 1-3)<br>5+ GCSE, O-levels, 1 A level, or equivalents (level 1-3)<br>2+ A levels or equivalents (level 1-3)<br>Undergraduate or postgraduate degree or other professional qualification (level 4)<br>Apprenticeship (none/other)<br>Other (e.g., vocational, foreign qualifications) (none/other)<br>Do not know (none/other)<br>Do not wish to answer (none/other) | level 1-3 |
|  | religious affiliation | atheist/agnostic<br>Christian<br>Buddhist (other religion)<br>Hindu<br>Muslim<br>other religion<br>do not wish to answer (not given) | atheist or agnostic |
|  | work status | working full-time (including self-employed)<br>part-time (including self-employed)<br>unemployed<br>student<br>looking after the home<br>retired (retired / disabled)<br>unable to work (e.g., short- or long-term disability) (retired / disabled)<br>do not wish to answer (other work status) | full-time |
|  | ethnicity | White: English/Welsh/Scottish/Northern Irish/British (White)<br>White: Irish (White)<br>White: Other white background (White)<br>White and Black Caribbean (mixed)<br>White and Black African (mixed)<br>White and Asian or White and Asian British (mixed)<br>Black, African, Caribbean or Black British (Black/Black British)<br>Asian or Asian British: Indian (Asian/Asian British)<br>Asian or Asian British: Pakistani (Asian/Asian British)<br>Asian or Asian British: Chinese (Asian/Asian British)<br>Asian or Asian British: Other (Asian/Asian British)<br>other ethnicity (other ethnicity)<br>do not wish to answer (other ethnicity) | White |
|  | language | English or Welsh<br>Polish<br>Punjabi (other language)<br>Urdu (other language)<br>Bengali (other language)<br>Other (other language)<br>do not wish to answer (other language) | English or Welsh |

In addition to the questions on baseline vaccination intent and change in vaccination inclination explained above, all respondents (n=16,527) are presented with a seven-item questionnaire to explore their attitudes towards vaccination and vaccine passports or certificates. These statements—which are answered on a scale from ‘*strongly agree*’ to ‘*strongly disagree*’ (with ‘*prefer not to say*’ a further option) and in which ‘social events’ are defined as above—are:

- Proof of vaccination via a vaccine certificate or passport for social events infringes on personal liberties
- I wish to be free to reject a vaccine without consequences on my ability to attend public or social events
- Individuals who reject a vaccine should not be allowed to attend social events
- Private companies should have the right to reject individuals if they have not received a vaccine
- Private companies should have the right not to employ unvaccinated staff
- Overall, I think vaccine passports are a good idea
- Requiring vaccine certificates or passports for social events is the same as requiring me to get vaccinated.

The order in which statements were presented to respondents was randomised.

### Estimating the impact of passports on vaccination inclination across the UK

To estimate the overall impact of the introduction of vaccine passports on intent to accept a COVID-19 vaccine nationally across the UK, we would like to estimate the distribution *P*_*Y,Z*_(*Y, Z*), where *Y* and *Z* denote the change in vaccination inclination if vaccine passports were introduced domestically (or internationally) and each individual’s baseline intent to accept a COVID-19 vaccine, respectively. This quantity gives the probability of each pair of responses (both modelled as ordinal random variables) and can be used to investigate how vaccine passports may shift vaccination intent. To estimate this quantity, we use multilevel regression and poststratification^23–25^ (MRP) to compute the posterior predictive distribution 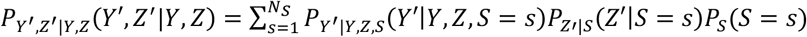 where *S* is an index variable that represents one of the *N*_*S*_ =370,440 unique census strata (12 regions × 2 sexes × 7 age groups × 3 education levels × 7 work statuses × 7 religious affiliations × 5 ethnicities × 3 primary languages). The first two terms in this equation are posterior predictive distributions obtained from ordinal multilevel logistic regressions which model the association between change in vaccination intention (*Y*) given existing vaccination intent (*Z*) and socio-demographic status and vaccination intent (*Z*) given socio-demographic status. The final term is a post-stratification step that re-weights these distributions according to the number of times a particular stratum appears in the UK census records.

To estimate the overall impact of vaccine passports/certificates on the UK publics’ inclination to accept a COVID-19 vaccine, we assume that respondents who state that they would ‘definitely’ accept or reject a COVID-19 vaccine will be no more or less inclined to vaccinate *unless* those replying ‘yes, definitely’ would *lower* their vaccination inclination if passports were introduced *or* if those replying ‘no, definitely not’ would *increase* their vaccination inclination. Thus, we can form the summary, *U*_*k*_ = *δ*_*k4*_ ∑_*j*>3_ 100 ∗ *P*_*Y′,Z′*_ (*Y′* = *j,Z′* = *k*) − *δ*_*k1*_ ∑_*j<*3_ 100 ∗ *P*_*Y′,Z′*_ (*Y′* = *j,Z′* = *k*), which measures the net shift in COVID-19 vaccination inclination induced by the introduction vaccine passports for each baseline intent level *k* (where *δ*_*i j*_ is Kronecker’s delta: *δ*_*i j*_ = 1 if *i* = *j* and 0 otherwise). This summary, in discounting individuals who already state a ‘definite’ intention to accept a COVID-19 vaccination and state that passports will increase their inclination to vote and, likewise, those who state a ‘definite’ intention not to accept a COVID-19 vaccination cannot be moved to a lower vaccination level, will capture the overall change in vaccination inclination relevant to baseline vaccination intent. The quantity *S* = ∑_*k*_*U*_*k*_ is a measure of the net population-wide possible change in vaccination intent pertinent to vaccination decisions induced by passports/certificates.

### Socio-demographic determinants of change in inclination to vaccinate

Ordinal multilevel logistic regressions are used to model *Y*|*Z, S* and *Z*|*S*, with separate models fitted for change in inclination *Y* given the introduction of passports for domestic and international use (see appendix). In addition, to assess the determinants of change in vaccination inclination without controlling for baseline intent, we also use ordinal multilevel regressions to calculate *Y*|*S*: this regression is not used in calculating the joint distribution *P*_*Y,Z*_(*Y, Z*) but illustrates the direct predictors of change in inclination without controlling for pre-existing intent. Fixed- and random-effect regression parameters from these models signify the association between socio-demographic status and change in vaccination intent induced by passports and intent to accept a COVID-19 vaccine at the national (fixed) and sub-national levels (random).

### Attitudes to vaccinations, passports, and societal freedoms

Seven individual MRP models are implemented to simultaneously estimate national (and sub-national) attitudes to the seven-item questionnaire and to explore their socio-demographic determinants. The response variable in each case are ordinal responses from each of the seven statements. Respondents who state that they ‘*prefer not to say’* are removed from each regression. A sensitivity analysis recoding ‘*prefer not to say*’ to ‘*neither agree nor disagree*’ is also performed to establish stability of estimates under a non-complete-case analysis (see appendix).

All multilevel regression models are implemented using JAGS version 4.3.0 (implemented via `rjags`^26^) and `R` version 4.0.3. 10,000 posterior samples (not including 2,000 for model burn-in) was sufficient for successful convergence and all posterior draws were well-mixed (see appendix). Post-stratification was implanted in `R` using UK census microdata (https://census.ukdataservice.ac.uk/get-data/microdata.aspx).

## Results

### UK-wide intent to accept a COVID-19 vaccine

Across the UK, we estimate that 78·71% (95% HPD interval, 76·14 to 81·04) of the adult population who have not yet had both doses of a COVID-19 vaccine will ‘definitely’ accept a future dose (either their first or second having already taken their first), while a further 11·72% (10·54 to 12·83) are ‘unsure but leaning towards yes’ (see figure 1 for sample information on doses received). 4·51% (3·84 to 5·24) say they will ‘definitely not’ accept a future COVID-19 vaccine, while 5·06% (4·43 to 5·73) are ‘unsure but leaning towards no’.

### Impact of vaccine passports on vaccine inclination

A large minority of respondents report that vaccination passports for domestic use (46·48%) or international travel (41·96%) would make them no more or less likely to accept a COVID-19 vaccine. Additionally, a sizeable minority of respondents also state that they would ‘definitely’ accept a COVID-19 vaccine and that vaccine passports would make them more likely to vaccinate (48·75% for domestic use and 42·89% for international travel), while 2·56% (2·32%) of respondents report that they would ‘definitely not’ accept a COVID-19 vaccine and that vaccine passports for domestic use (international travel) would make them less likely to vaccinate (table 2).

**Table 2.** Change in vaccination inclination if vaccine passports or certificates introduced for domestic and international use is strongly associated with existing vaccination intent.

| $100 * P_{Y',Z'\vert Y,Z}(Y', Z' \vert Y, Z)$ | | Existing vaccination intent, $Z'$ | | | |
| --- | --- | --- | --- | --- | --- |
|  |  | No, definitely not (1) | Unsure, but leaning towards no (2) | Unsure, but leaning towards yes (3) | Yes, definitely (4) |
| **Change in inclination to accept vaccine if passports introduced for domestic use, $Y'$** | Much less likely (1) | 1.88 [1.57, 2.21] | 1.03 [0.87, 1.21] | 0.87 [0.73, 1.02] | 2.62 [2.21, 3.05] |
|  | Somewhat less likely (2) | 0.68 [0.58, 0.80] | 0.59 [0.52, 0.68] | 0.66 [0.57, 0.76] | 2.15 [1.84, 2.47] |
|  | Neither more nor less likely (3) | 1.74 [1.43, 2.05] | 2.84 [2.47, 3.22] | 6.71 [6.07, 7.36] | 35.19 [32.60, 37.82] |
|  | Somewhat more likely (4) | 0.10 [0.08, 0.12] | 0.29 [0.23, 0.34] | 1.45 [1.27, 1.65] | 13.10 [12.55, 13.59] |
|  | Much more likely (5) | 0.10 [0.08, 0.12] | 0.31 [0.25, 0.38] | 2.03 [1.67, 2.35] | 25.65 [23.12, 28.33] |
| **Change in inclination to accept vaccine if passports introduced for international use, $Y'$** | Much less likely (1) | 1.62 [1.33, 1.89] | 0.77 [0.65, 0.90] | 0.73 [0.62, 0.85] | 2.49 [2.13, 2.82] |
|  | Somewhat less likely (2) | 0.70 [0.59, 0.80] | 0.51 [0.44, 0.59] | 0.59 [0.51, 0.68] | 2.14 [1.87, 2.42] |
|  | Neither more nor less likely (3) | 1.90 [1.61, 2.23] | 2.85 [2.51, 3.24] | 6.03 [5.44, 6.68] | 31.18 [28.92, 33.24] |
|  | Somewhat more likely (4) | 0.14 [0.12, 0.17] | 0.41 [0.35, 0.47] | 1.61 [1.43, 1.78] | 12.72 [12.27, 13.12] |
|  | Much more likely (5) | 0.16 [0.13, 0.19] | 0.52 [0.44, 0.62] | 2.76 [2.4, 3.14] | 30.17 [27.70, 33.02] |

In assessing the impact of vaccine passports on vaccine intention, however, it is essential to consider individuals for whom vaccine passports will likely alter their ultimate decision to vaccinate. The summary metric *U*_*k*_ excludes these individuals and suggests that vaccine passports may result in a *lower* overall inclination to accept COVID-19 vaccines. Overall, while vaccine passports for domestic use may have a very small positive impact on those who report that they will ‘definitely not’ accept a COVID-19 (*U*_1_=0·20, 95% HPD interval 0·15 to 0·24), they will likely have a negative impact on those who report that they would otherwise have ‘definitely’ accepted a COVID-19 vaccine (*U*_1_=-4·77, -5·53 to -4·05). The overall net impact *S* = ∑_*k*_*U*_*k*_ suggests a loss of intent to vaccinate of -3·64, -5·26 to -2·06 (table 3). Similar results are found for the impact of vaccine passports for international travel; however, the overall net loss of intent is lower, with *S* = -1·65 (−2·97 to -0·06), suggesting that vaccine passports for international travel are less disagreeable than for domestic purposes, with *S*^DOM^ - *S*^INT^ =-1·99 (−3·87 to 0·13).

**Table 3.** Potential impact of passports on inclination to receive COVID-19 vaccine.

| | Existing vaccination intent, $Z'$ | | | | $S = \sum_k U_k$ |
| --- | --- | --- | --- | --- | --- |
|  | No, definitely not (1) | Unsure, but leaning towards no (2) | Unsure, but leaning towards yes (3) | Yes, definitely (4) |  |
| **Overall impact if passports introduced for domestic use**<br>$U_k^{\text{DOM}}$ | 0.20 [0.15, 0.24] | -1.03 [-1.30, -0.77] | 1.95 [1.28, 2.54] | -4.77 [-5.53, -4.05] | -3.64 [-5.26, -2.06] |
| **Overall impact if passports introduced for international use, $U_k^{\text{INT}}$** | 0.30 [0.24, 0.36] | -0.36 [-0.58, -0.11] | 3.05 [2.49, 3.66] | -4.64 [-5.27, -4.03] | -1.65 [-2.97, -0.06] |
| **Difference in impact effect size between international and domestic use, $\Delta S = S^{\text{DOM}} - S^{\text{INT}}$** | -0.10 [-0.14, -0.05] | -0.67 [-1.02, -0.36] | -1.09 [-1.83, -0.31] | -0.12 [-1.01, 0.82] | -1.99 [-3.87, 0.13] |

The socio-demographic determinants of self-reported change in vaccination inclination if passports are introduced for domestic use are shown in figure 2 while controlling for baseline vaccination intent, *Y*|*Z, S* (figure 2A) and without this control, *Y*|*Z* (figure 2B). Similarly, these two sets of determinants for international travel are shown in figure 3. We interpret odds ratios for which the corresponding 95% highest posterior density interval excludes zero as ‘significant’.

**Figure 2.**
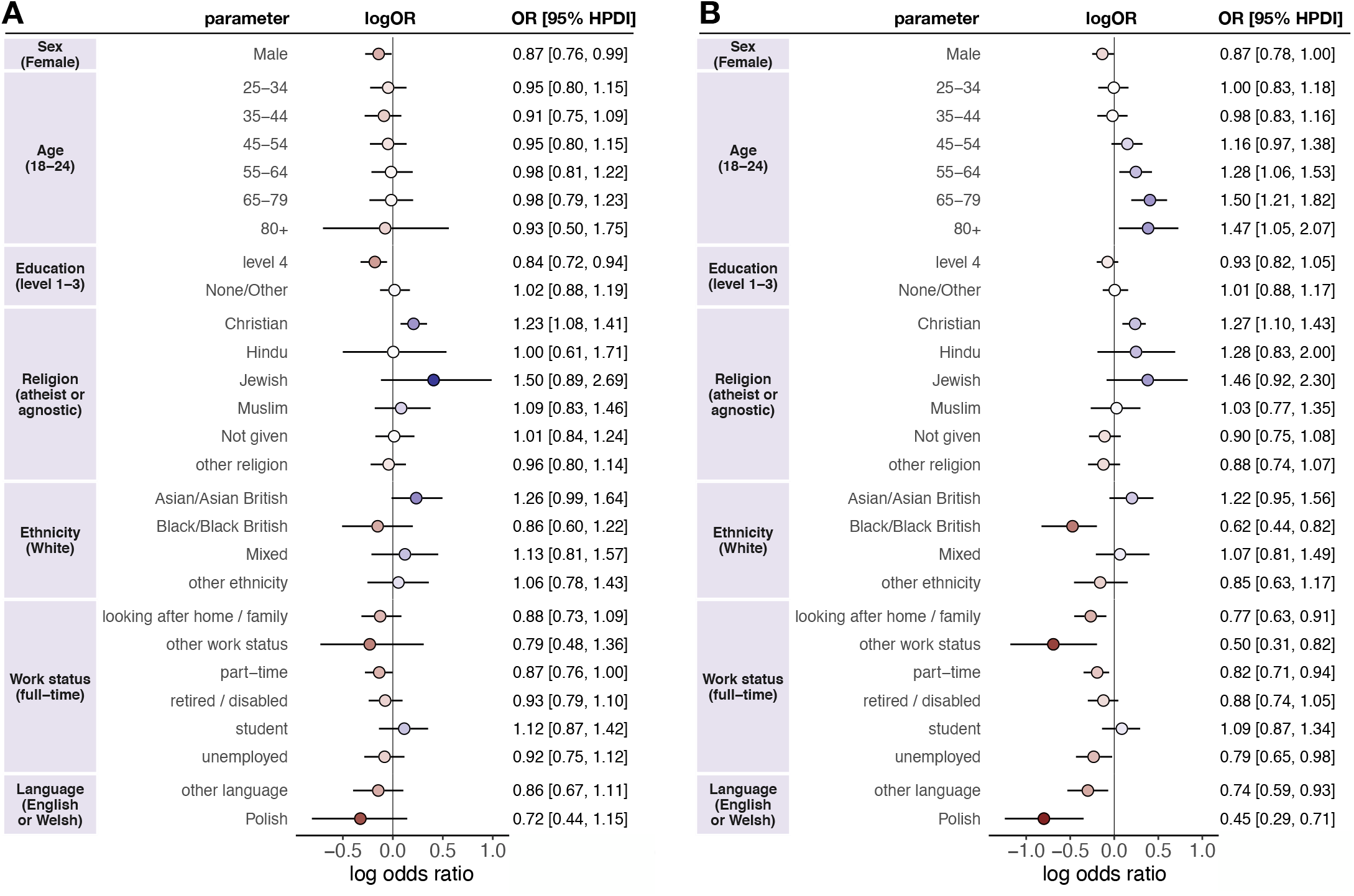
**Socio-demographic determinants of change in vaccination inclination**, *Y*, **if vaccine passports were required for domestic use with a control for baseline vaccination intent**, *Z* **(A) or no control (B)**. Multilevel regression fixed-effect parameter log odds ratios are plotted with corresponding 95% highest posterior density intervals. Baseline intent to accept a vaccine, *Z*, is not shown in A for visual purposes, but the log odds ratio is 3·11 (2·87 to 3·30): this parameter is denoted $_*Z*_ in the model formulation (appendix). Log odds ratios are coloured by effect magnitude and direction, where blues (reds) signify that the group is more (less) inclined than the baseline group to accept a COVID-19 vaccine and the darker the colour the stronger the association. For each factor, the baseline group is provided in parentheses on the left. Odds ratios with 95% HPDIs are shown on the right for each parameter.

**Figure 3.**
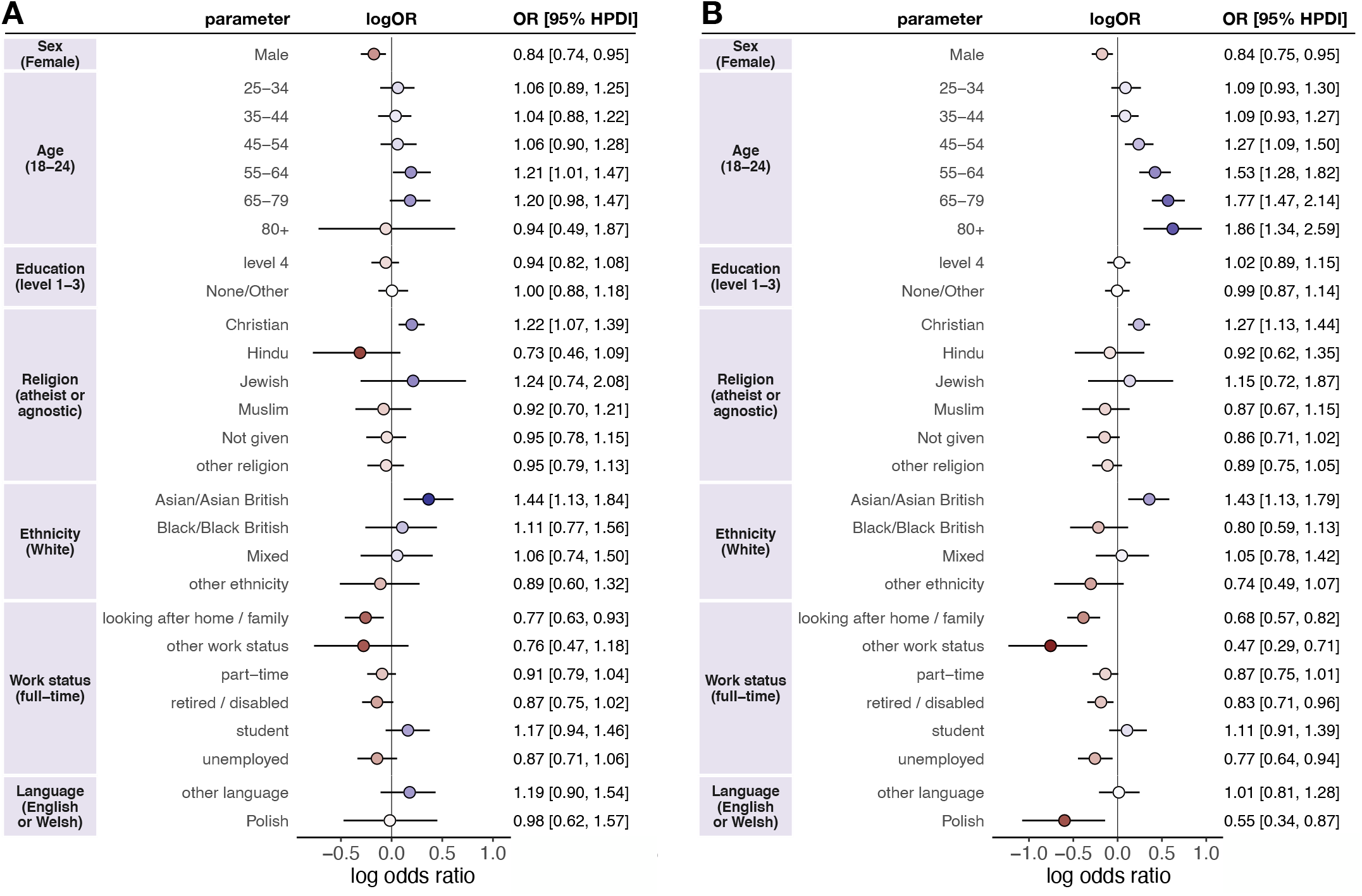
**Socio-demographic determinants of change in vaccination inclination**, *Y*, **if vaccine passports were required for international travel with a control for baseline vaccination intent**, *Z* **(A) or no control (B)**. Multilevel regression fixed-effect parameter log odds ratios are plotted with corresponding 95% highest posterior density intervals. Baseline intent to accept a vaccine, *Z*, is not shown in A for visual purposes, but the log odds ratio is 2·90 (2·70 to 3·09): this parameter is denoted $_*Z*_ in the model formulation (appendix). Log odds ratios are coloured by effect magnitude and direction, where blues (reds) signify that the group is more (less) inclined than the baseline group to accept a COVID-19 vaccine and the darker the colour the stronger the association. For each factor, the baseline group is provided in parentheses on the left. Odds ratios with 95% HPDIs are shown on the right for each parameter.

Males (odds ratio, OR, 0·87, 95% highest posterior density interval—HPDI—0·78 to 1·00), Black and Black British ethnicities (0·62, 0·44 to 0·82), those unemployed (0·79, 0·65 to 0·98), looking after the home or family (0·77, 0·63 to 0·91), with in part-time employment (0·82, 0·71 to 0·94), or who have another work status (0·50, 0·31 to 0·82), or who speak Polish (0·45, 0·29 to 0·71) or another language (0·74, 0·59 to 0·93) report that they would be less inclined to vaccinate if passports were introduced for domestic use (compared to females, whites, and those in full-time employment, respectively) (figure 2B). Age groups above 45-55 and Christians (1·27, 1·10 to 1·43) report that they would me more inclined to vaccinate than 18-24-year-olds and atheists or agnostics. After controlling for baseline vaccination intent, males (0·87, 0·76 to 0·99) and those with undergraduate/postgraduate degrees or other professional qualifications (level 4) (OR 0·84, 0·72 to 0·94) are less inclined to accept a COVID-19 vaccine if vaccine passports are introduced for domestic use than females or those with level 1-3 education (undergraduate or postgraduate degrees, see table 1), respectively, while Christians (OR 1·23, 1·08 to 1·41) are more inclined than atheists or agnostics (figure 2A). There is also an extremely strong association between change in vaccination inclination and baseline vaccination intent (OR 3·11, 2·87 to 3·30, figure 2A). There is sub-national variability around these national level (fixed effects) estimates. For instance, respondents identifying as Asian or Asian British state more inclination (than Whites) to accept COVID-19 vaccines if passports were introduced for domestic use in West Midlands and London, while Jewish respondents in London state much more inclination than atheists or agnostics (table 2, appendix). Those unemployed in Yorkshire and the Humber, individuals working part-time in London, and retired / disabled respondents in South West England are less inclined to vaccinate than those in full-time employment. While students in London are much more inclined to vaccinate if vaccine passports were introduced for domestic use (table 2, appendix).

Similar trends are observed for determinants of change vaccination inclination if passports were introduced for international use (figure 3B). Males, those unemployed, looking after the home or family or with another work status, and Polish speakers still report being less inclined (than females, those in full-time employment, and English or Welsh speakers) to vaccinate if passports were introduced for international travel. Those retired or disabled (0·83, 0·71 to 0·96) now report being less inclined compared to those in full-time employment, while Black ethnicities are now no more or less likely than whites. Older age groups, Christians, and now Asian or Asian British ethnicities report being more inclined to vaccinate than 18-24-year-olds, atheists or agnostics (figure 3B). After controlling for baseline vaccination intent, males (0·84, 0·74 to 0·95) and those looking after the home or family (OR 0·77, 0·63 to 0·93) are less inclined to accept a COVID-19 vaccine if vaccine passports are introduced for international travel use than males or those in full-time employment, respectively; while 55-64-year-olds (OR 1·21, 1·01 to 1·47), Christians (OR 1·22, 1·07 to 1·39), and Asian or Asian British ethnicities (OR 1·44, 1·13 to 1·84) are more inclined than 18-24-year-olds, atheists or agnostics, or Whites, respectively (figure 3A). There is again an extremely strong association between change in vaccination inclination and baseline vaccination intent (OR 2·90, 2·70 to 3·09, figure 3A). Sub-national socio-demographic trends are shown in appendix, table 3: individuals who report looking after the home or family are considerably less likely than those in full-time employment to be inclined to vaccinate if passports were introduced for international travel in four UK regions, while students in London are again more inclined.

### Attitudes to vaccinations, passports, and societal freedoms

Overall, a majority of the UK public yet to have both doses believe that vaccination certificates or passports is the same as requiring vaccination (58·4%, 56·8 to 62·6), yet a majority of the public believe that they are a good idea (59·8%, 56·8 to 62·6). More respondents believe that passports do not infringe on personal liberties (41·1%, 38·2 to 43·8) than do (35·5%, 32·9 to 38·83), though this difference is smaller than in the two statements above. More respondents also state that they do not wish to be free to reject a vaccination without consequences on their ability to attend public or social events (39·3%, 36·6 to 42·2 disagree versus 33·2%, 30·7 to 35·8 who agree). A small majority of respondents also believe that individuals who reject a vaccine should not be able to attend social events (50·8%, 47·4 to 54·0). A breakdown of all seven statements, including views on the rights of private companies (not commented on here) can be found in appendix, figure 2.

There is a consistency in how many socio-demographic groups reply across these statements (appendix, figure 3), which also reflects responses to both change in vaccination inclination (figure 2B and figure 3B) and baseline vaccination intention (appendix, figure 1). For example, older age groups—who state a higher intent to vaccinate and also an increased inclination to vaccinate if passports were introduced—are more likely to agree that passports are a good idea; that passports would not infringe their personal liberties; that they do not wish to be free to reject a vaccine; and that individuals who reject a vaccine should not be free to attend social events. However, Black and Black British respondents are more likely than Whites to believe that vaccine passports would infringe on their personal liberty; that they wish to be free to reject a vaccine without consequences on their ability to attend social events. Black and Black British respondents are also much less likely than Whites to think that individuals who reject a vaccine should be allowed to attend social events and are less likely than Whites to think vaccine passports are a good idea. A full set of regression parameters for each statement is provided in appendix, figure 3.

## Discussion

The overall data from our study suggest a somewhat reassuring picture. Vaccine passports have a positive impact on stated intentions to get vaccinated amongst those who have not received doses. However, in practical terms, these are not the data that concern us.

There are two issues that this study raises that are critical to any future use of vaccine passports in the UK. First, at an individual level, it is of little help to make those who have already made a definite decision to get vaccinated feel even more positive about doing so (and these comprise nearly 80% of respondents). The important question is what effect will such passports have on those who are having doubts and could be swayed one way or another? Second, at a collective level, the levels of uptake in some communities are high; however, there are some regions where uptake is much lower^27^. These areas are typified by larger populations of younger age groups, non-White ethnicities, and non-English speakers^25,27^. As we find in this study, vaccine passports or certificates are viewed less positively within these socio-demographic groups: this creates a risk of creating a divided society wherein the majority are relatively secure but there remain pockets of lower vaccination where outbreaks can still occur. This latter point is especially important in the context of local vaccination rates required to prevent epidemic spread. The important question here concerns the impact of vaccine passports amongst members of these communities: are they likely to help or hinder efforts to ensure that overall levels of immunity are uniformly high amongst all sections of the population? If we cannot persuade groups in localised clusters to get vaccinated—or worse, enact policies which may lower their confidence in vaccines—then these areas are not only at increased epidemic risk, but may serve to increase required vaccination levels for herd immunity in adjacent settings^28^.

In regard to the first issue, when we remove those participants who express certainty (they either definitely will or definitely will not get a jab) and an unchanged inclination to vaccinate were passports introduced, and focus on the remaining doubters, a very different picture emerges. Overall, this remaining group expresses *lower* intentions to get vaccinated when vaccine passports are mentioned, especially when these passports cover domestic activities as opposed to international travel. Recent evidence among health and social care workers in the UK suggests that feeling pressured to vaccinate is associated with lower uptake^29^.

Turning to the second issue, when we break our overall dataset down and look at the effects of vaccine passports on vaccination intentions upon different groups, we find considerable variability. While males have been reported to have a higher intent to accept COVID-19 vaccines in the UK than females^14,30,31^ (which is being borne out across most age groups who have been offered COVID-19 vaccines—see appendix, table 4), we find that, after controlling for baseline vaccination intent, it is males who are more likely to lower their vaccination inclination if vaccine passports are introduced, accounting for baseline vaccination views. Similarly, those with university degrees and other professional qualifications also report a decreased inclination to vaccinate (compared to level 1-3 education, see figure 2 and table 1 for variable definition) if vaccine passports are introduced for domestic use. There is notable sub-national variation in these trends, for example Jewish respondents in London, passports (both for international and domestic use) increase stated vaccination inclination. It is also notable that, among the groups with lower observed uptake—such as the Black community and those who are economically deprived (unemployed)—the effects of domestic use vaccine passports on stated vaccination inclination (without controlling for baseline intent) are most negative.

As to why we get such different responses in different groups, it is impossible to be certain. However, based on research into the ways that the public make sense of new and unfamiliar scientific phenomena by assimilating them to prior and familiar schemata, one key place to look for an answer is in the shared beliefs (or ‘social representations’) of the relevant groups^32^. In the case of Jewish groups, of whom many may identify with Israel, the positive view of passports may relate to the publicity given to the ‘green pass’ system in Israel^33^, however, some Israeli medical professors have recently cited segregation of those vaccinated from those unvaccinated by universities and public venues, as well as vaccine rejection by younger Israelis who would “have never considered refusing a vaccine recommended to them”^34^. By contrast, amongst the Black community, the negative impact of passports may relate to a longstanding suspicion, buttressed by historical experience^35^, that medical interventions are used as a means of controlling the community^36^. In other words, Black people are more likely to see vaccine passports—especially when they impinge on everyday activities—as something imposed on them rather than something provided for them and therefore are put off vaccination when they are invoked. Our data reveal that Black respondents are more likely than White respondents to believe that vaccine passports are an attack on civil liberties and are in less agreement that people who reject vaccines should be disqualified from social events. However, we cannot say from our findings that there is any causal link between historically grounded perceptions of compulsion and control on the one hand and lowered vaccine intentions on the other. Clearly, this is a key site for further investigation.

Before drawing any conclusions from our study, it is important to note a number of limitations. Most obviously, our data consists of self-reports and, due to social desirability concerns, people may over-state their intention to do what the Government, health professions and media are heavily promoting. However, comparing those who either said they would definitely get vaccinated or were leaning towards it in a nationally representative survey of 16,820 UK adults conducted in October 2020^14^ (two months before the first COVID-19 vaccination in the UK^37^) to the numbers of people who actually took up the offer of a first dose (to 30 April 2021), we find that the former is close to but actually a little lower than the latter (55-64-year-olds: 83·8% vs. 86·9%; 65-79-year-olds: 90·0% vs. 93·2%; Over 80s: 90·1% vs. 94·9%). In sum, without ruling it out, there is little evidence for a strong desirability effect, although previous underestimation could also point to a general increase in intent to vaccinate across the UK since vaccine rollout began.

A second and related issue is that, even if it is accepted that our data reflect genuine intentions, these are still not the same as actual vaccination uptake. While we find that passports result in a net decrease in vaccination inclinations among those who are undecided, we have no way of knowing whether this is enough to tip anybody over into actually refusing the vaccine. Hence, we cannot be definitive about the real-world impact of introducing such passports.

Finally, it is important to stress that the discussion of COVID passes and passports covers many different possibilities which vary along at least three dimensions: what it depends on (vaccination, negative PCR/lateral flow test results, antibody testing); what it applies to (international travel, attendance at large events, access to pubs/restaurants, shops, employment); when it applies (immediately, after everyone has been offered vaccination). We suspect that the impact of passport proposals on vaccination intentions will vary as a function of all of these factors (which will impact on possible mediating process such as perceptions of compulsion or else perceptions of legitimacy and equity).

We also acknowledge that the alternatives included in our study (1. passports for international travel; 2. Passports for social events including sports events, bars and restaurants) are not necessarily the alternatives offered in any particular country (for instance, in the UK currently proposals are being mooted to introduce vaccine passports for sports events but not for pubs and restaurants^38^). Our precise findings may not be directly applicable, therefore. However, our more general message remains highly relevant across different settings. Moreover, we have not considered whether, for example, *not* introducing passports to facilitate international travel may disincentivise vaccine uptake among those who have stated an intent to vaccinate, especially if financial costs are incurred to prove immunological status to travel.

What we have described is what might be dubbed a ‘vaccine passport paradox’ whereby the overall positivity of a population towards the introduction of passports may mask processes that alienate critical minorities and may possibly lead to an overall decrease in inclination to vaccinate. This is not an argument against vaccine passports in general. There may be some variants of passport schemes which do not create reactance in these critical minorities. But it is to introduce a note of caution to the debate. Before making decisions on the introduction of any specific vaccine passport policy, it is necessary to address the impact of passports on the decisions of those individuals and communities who are more hesitant about vaccination and hence most need to be persuaded to take them.

This study has implications for the UK’s policy on vaccine certification as well as, more broadly, implications for other countries who are planning to introduce vaccine certification. For example, a European Union digital green certificate is being created with the purpose to facilitate free movement inside the EU during the COVID-19 pandemic^2^. However, it is currently unclear whether such a pass could result in lower inclination to vaccinate across European Union member states, notably those who have expressed concerns over the safety and importance of vaccines in recent years^39^. As passports are unpopular with groups with low intent to vaccinate, the introduction of passports could have profound consequences in settings where there are lower levels of baseline confidence in vaccines, such as France and Poland^40^.

In conclusion, our study suggests that vaccine passports may induce a lower vaccination inclination in socio-demographic groups that are less confident in COVID-19 vaccines. As these groups tend to cluster geographically in large urban areas, extreme caution should be exercised in any public health intervention that may lead to less positive health-seeking behaviours in areas at high epidemic-risk.

## Data Availability

Data is available upon request to the corresponding author.

## Ethical approval

Ethical approval for this study was obtained via the London School of Hygiene and Tropical Medicine’s Research Ethics Committee on 7 April 2021 with reference 25637 and, for October 2020 survey data, the Imperial College Research Ethics Committee on 24 July 2020 with reference 20IC6133.

