## appendix for "The potential impact of vaccine passports on inclination to accept COVID-19 vaccinations in the United Kingdom: evidence from a large cross-sectional survey and modelling study"

### Supplementary methods

#### Multilevel regression and poststratification to estimate $P_{Y,Z}(Y,Z)$ and establish socio-demographic determinants of $Y|Z$ and $Z$

We wish to estimate the joint distribution $P_{Y,Z}(Y,Z)$, where $Y$ denotes change in vaccination inclination if vaccine passports were introduced for use domestically for social events (or, using the same notation but an independent model implementation, for international use) and $Z$ denotes each individuals' baseline intent to accept a COVID-19 vaccine. Both $Y$ and $Z$ are modelled as ordinal random variables. We want to obtain the posterior predictive distribution $P_{Y\prime,Z'|Y,Z}(Y',Z'|Y,Z) = \mathbb{E}_S P_{Y\prime|Y,Z,S}(Y'|Y,Z,S)P_{Z\prime|X}(Z'|S)$, where $S$ is an index variable that represents one of the $N_S$ =370,440 unique census strata (12 regions × 2 sexes × 7 age groups × 3 education levels × 7 work statuses × 7 religious affiliations × 5 ethnicities × 3 primary languages). Thus,

$$P_{Y\prime,Z\prime}(Y',Z'|Y,Z) = \sum_{s=1}^{N_S} P_{Y\prime|Y,Z,S}(Y'|Y,Z,S=s)P_{Z\prime|X}(Z'|S=s)P_S(S=s). \quad (1)$$

The first two elements on the right-hand side of equation (1) are estimated via posterior predictive distributions,

$$P_{Y\prime|Y,Z,S}(Y'|\,Y,Z,S) = \int P_{Y|Z,S,\theta}(Y'|Z,S,\theta)P_{\theta|Z,S,Y}(\theta|Z,S,Y)\mathrm{d}\theta \quad (2)$$

$$P_{Z\prime|Z,S}(Z'|\,Z,S) = \int P_{Z\prime|Z,S}\big(Z'\big|Z,S,\tilde{\theta}\big)P_{\tilde{\theta}|Z,S}(\tilde{\theta}|Z,Y)\mathrm{d}\tilde{\theta} \quad (3)$$

obtained from two separate multilevel regressions, where both regression models deploy ordinal logistic regressions (see below) and where $P_{\theta|Z,S,Y}(\theta|Z,S,Y)$ and $P_{\tilde{\theta}|Z,S}(\tilde{\theta}|Z,Y)$ denote posterior distributions over all model parameters $\theta$. The last term of equation (1) simply counts the number of unique strata in the UK census records and divides by the total number of individuals, $P_S(S=s) = \#(S=s)/N$. That is, this final term implements post-stratification according to UK census microdata records.

Multilevel ordinal logistic regression is used to model $Y|Z,S$,

$$Y_j(i)|Z_j(i),S_j(i) \sim \mathrm{OrderedLogistic}(\beta_Z \sum_{i=1}^{Z_j(i)-1} \delta_{ij} + f\big(S_j(i);\, \boldsymbol{\beta}_j\big);\, \tau_1,\tau_2,\tau_3,\tau_4) \quad (5)$$

where $Y_j(i) \in \{1,2,3,4,5\}$ denotes the change in inclination for individual $i$ in sub-national region $j$ (where, as a reminder, 1=much less likely and 5=much more likely, see main text table 1); $S_j(i)$ denotes individuals' socio-demographic characteristics; the term $\beta_Z \sum_{i=1}^{Z_j(i)-1} \delta_{ij}$ models that $Z_j(i) \in \{1,2,3,4\}$ is an ordinal covariate[41]; and where $\delta_{ij} \geq 0$ and $\sum_{i=1}^{4} \delta_{ij} = 1\, \forall j$, $-\infty < \tau_1 < \tau_2 < \tau_3 < \tau_4 < \infty$. We use the ordered logistic distribution for $k$ ordered outcomes specified by $Y \sim \mathrm{OrderedLogistic}(\beta; \tau_1, \tau_2, \dots, \tau_{k-1})$, where $P_Y(Y \leq k) = \sigma(\tau_k - \beta)$ and $\sigma(x) = 1/(1+e^{-x})$ is the standard logistic sigmoid function. This definition is equivalent to the proportional-odds assumption, wherein the difference in the log of cumulative odds ratios between successive categories is independent of the slope $\beta$. The function $f\big(S_j(i);\, \boldsymbol{\beta}_j\big) = \beta_{sex(i),j} + \beta_{age(i),j} + \beta_{edu(i),j} + \beta_{rel(i),j} + \beta_{wrk(i),j} + \beta_{eth(i),j} + \beta_{lan(i),j}$, where $\forall\, d \in D = \{sex, age, edu, rel, wrk, eth, lan\}$, $d(i)$ corresponds to the category to which $i$ belongs and $\beta_{d(i)} = 0$ if $d(i)$ is a baseline group (male, aged 18-24, with level 1-3 education, a Christian, in full-time employment, White ethnicity, and speaks English/Welsh/Scottish as a first language—see main text table 1 for full definitions).

Regularising priors are placed on all socio-demographic fixed-effect parameters $\beta_{k,j} \sim N(\gamma_k, p_k)$ (where $p_k = \frac{1}{\sigma_k^2}$ is the precision) and $p_k \sim Gamma(1,.1)$ places a relatively uniform prior over corresponding values for $\sigma_i > 1$, with less prior mass assigned as $\sigma_i$ decreases from 1 towards 0. (Here $k$ indexes over all random-effect parameters $d$.) $\gamma_k \sim N(0, 10^{-1}) \forall k$, which places a semi-informative prior over fixed-effect parameters with variance equal to 10.

In addition, the second term in equation (1) is modelled as,

$$Z_j(i)|\, S_j(i) \sim OrderedLogistic(f\big(S_j(i);\, \boldsymbol{\beta}'_j\big);\, \tau'_1, \tau'_2, \tau'_3, \tau'_4), \quad (6)$$

with the same conditions on the $\tau$ parameters in the ordered logistic function as specified above and with $f\big(S_j(i);\, \boldsymbol{\beta}'_j\big) = \beta'_{sex(i),j} + \beta'_{age(i),j} + \beta'_{edu(i),j} + \beta'_{rel(i),j} + \beta'_{wrk(i),j} + \beta'_{eth(i),j} + \beta'_{lan(i),j}$. This multilevel ordinal logistic regression estimates the relationship between socio-demographic factors and baseline intent to accept a COVID-19 vaccine. The prime notation distinguishes the parameters from those in equation (5). The same prior distributions are used as for the fixed- and random-effect parameters for equation (5).

We also estimate the determinants of change in vaccination inclination directly (that is, without controlling for baseline vaccination intent) via,

$$Y_j(i) | \, S_j(i) \sim OrderedLogistic(f(S_j(i);\, \boldsymbol{\beta}''_j);\, \tau''_1, \tau''_2, \tau''_3, \tau''_4), \quad (7)$$

where all parameters and respective priors are as defined above, but the double primed notation differentiates parameters between models.

##### Multilevel regression and poststratification to estimate responses to the seven-item battery on vaccinations and societal freedoms

Multilevel regression and poststratification is also used to estimate national-level sentiment and the socio-demographic determinants of the seven questions outlined in the main text. Seven individual MRP models are implemented to simultaneously estimate national attitudes to these statements and to explore their socio-demographic determinants. These models follow the same process as per equation (1), but instead we estimate $P_{Z_q'}(Z_q' | Z_q)$, where $Z_q$ denotes the response to one of the seven statements: (1) Proof of vaccination via a vaccine certificate or passport for social events infringes on personal liberties; (2) I wish to be free to reject a vaccine without consequences on my ability to attend public or social events; (3) Individuals who reject a vaccine should not be allowed to attend social events; (4) Private companies should have the right to reject individuals if they have not received a vaccine; (5) Private companies should have the right not to employ unvaccinated staff; (6) Overall, I think vaccine passports are a good idea; (7) Requiring vaccine certificates or passports for social events is the same as requiring me to get vaccinated.

##### Estimating age-stratified intention to accept a COVID-19 vaccine using data from October 2020

In October 2020, a nationally representative survey of 16,820 UK adults conducted in October 2020 asked respondents '*If a new coronavirus (COVID-19) vaccine became available, would you accept the vaccine for yourself?*' with the same four possible responses as provided in this study (that is, '*yes, definitely*', '*unsure, but leaning towards yes*, '*unsure, but leaning towards no*, and '*no, definitely not*'). These data are used in the main text discussion to assess whether age-specific intent recorded in October 2020 matched uptake among individuals over 55, to thus establish whether a possible key limitation of the study—social desirability biases—may play a strong limiting factor in our analysis.

Intent to accept a COVID-19 vaccine in October 2020 in the UK is estimated for each of three birth cohorts who have all been offered a COVID-19 vaccine at the time of the study. These birth cohorts are: 55-64 (n=2,059), 65-79 (n=2,623), and 80+ year-olds (n=157). Uptake intent (as measured in October 2020) is estimated using the model, $\boldsymbol{Y}_c \sim Multinom(\boldsymbol{\pi}_c, n_c)$, with $\boldsymbol{\pi}_c \sim Dirichlet(1,1,1,1)$, where $\boldsymbol{Y} = (Y_1, Y_2, Y_3, Y_4)$ is the number of respondents falling into each of the four categories (1 = '*No, definitely not*', 2='*Unsure, but leaning towards no*', 3='*Unsure, but leaning towards yes*', 4='*Yes, definitely*').

### Supplementary figures and tables

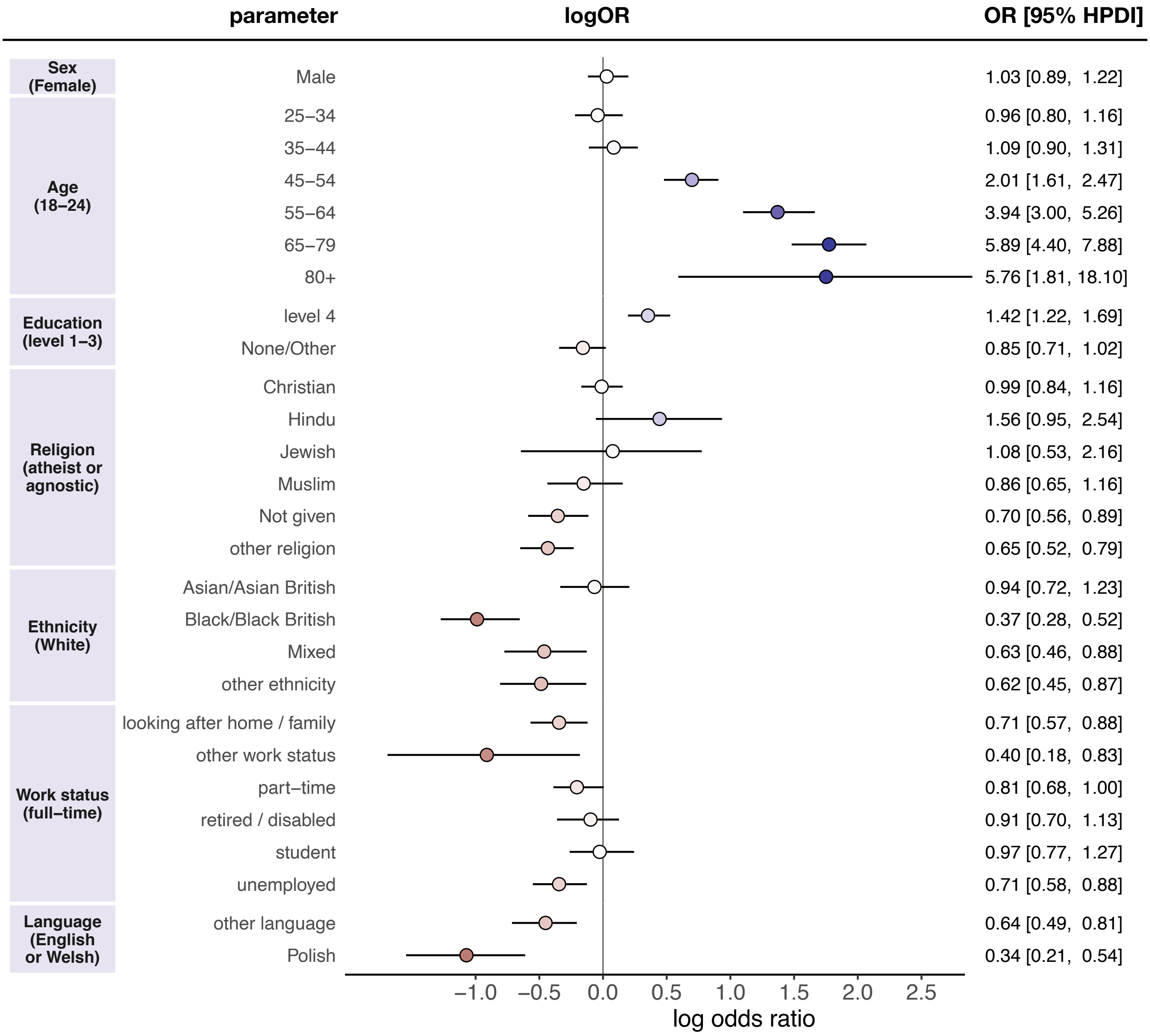


**Figure 1 | Socio-demographic determinants of baseline vaccination intent,** $z$. Multilevel regression fixed-effect parameter log odds ratios are plotted with corresponding 95% highest posterior density intervals. Log odds ratios are coloured by effect magnitude and direction, where blues (reds) signify that the group is more (less) inclined than the baseline group to accept a COVID-19 vaccine and the darker the colour the stronger the association. For each factor, the baseline group is provided in parentheses on the left. Odds ratios with 95% HPDIs are shown on the right for each parameter.

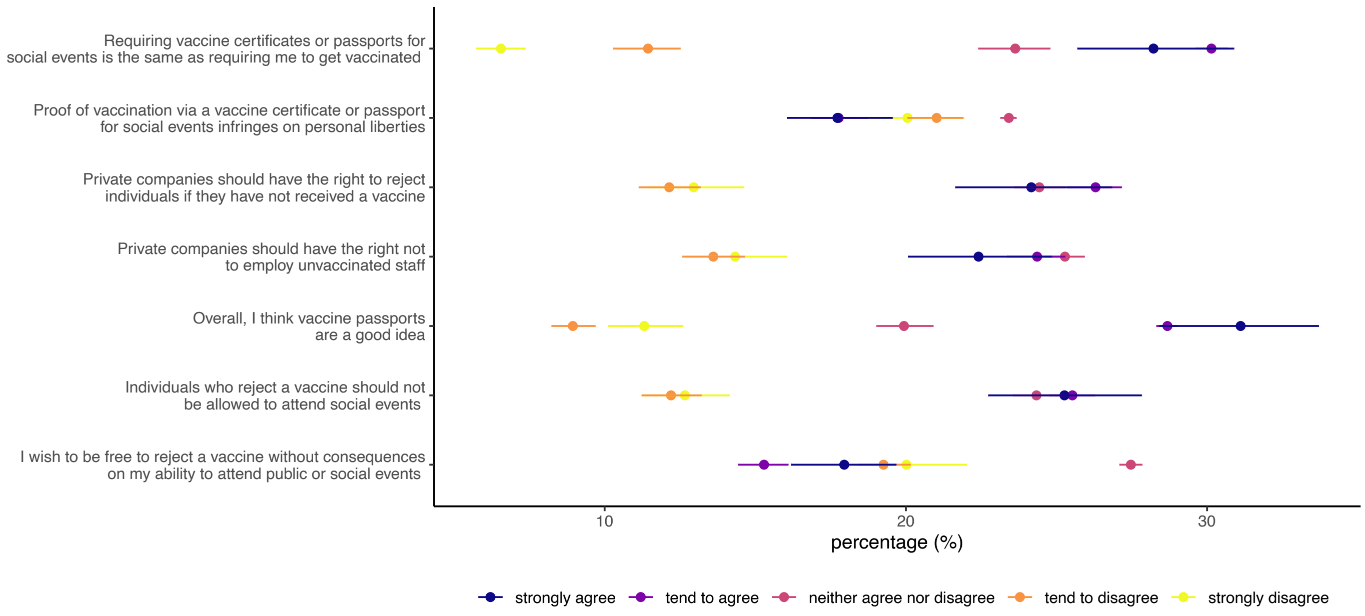


**Figure 2 | MRP estimates of seven-item battery on attitudes to vaccinations, passports, and societal freedoms** Percentage of the UK population who have not yet received both doses who agree to seven statements on vaccinations, passports, and societal freedoms. Horizontal bars denote 95% highest posterior density intervals. Overall agreement (disagreement)—that is the sum of 'strongly agree' ('strongly disagree') and 'tend to agree' ('tend to disagree')—to the seven statements is as follows: Requiring vaccine certificates or passports for social events is the same as requiring me to get vaccinated (agree: 35.5% [32.9 to 38.3]; disagree: 41.1% [38.2 to 43.8]); I wish to be free to reject a vaccine without consequences on my ability to attend public or social events (agree: 33.2% [30.7 to 35.8]; disagree: 39.3% [36.6 to 42.1); Individuals who reject a vaccine should not be allowed to attend social events (agree: 50.8% [47.4 to 54.0]; disagree: 24.9% [22.4 to 27.3]; Private companies should have the right to reject individuals if they have not received a vaccine (agree: 50.5% [47.0 to 53.9]; disagree: 25.1% [22.6 to 27.8]; Private companies should have the right not to employ unvaccinated staff (agree: 46.8% [43.5 to 50.11]; disagree: 27.9% [25.3 to 30.7]); Overall, I think vaccine passports are a good idea (agree: 59.8% [56.8 to 62.6]; disagree: 20.3% [18.3 to 22.3]); Requiring vaccine certificates or passports for social events is the same as requiring me to get vaccinated (agree 58.4% [55.3 to 61.6]; disagree 18.0% [16.0 to 19.9]).

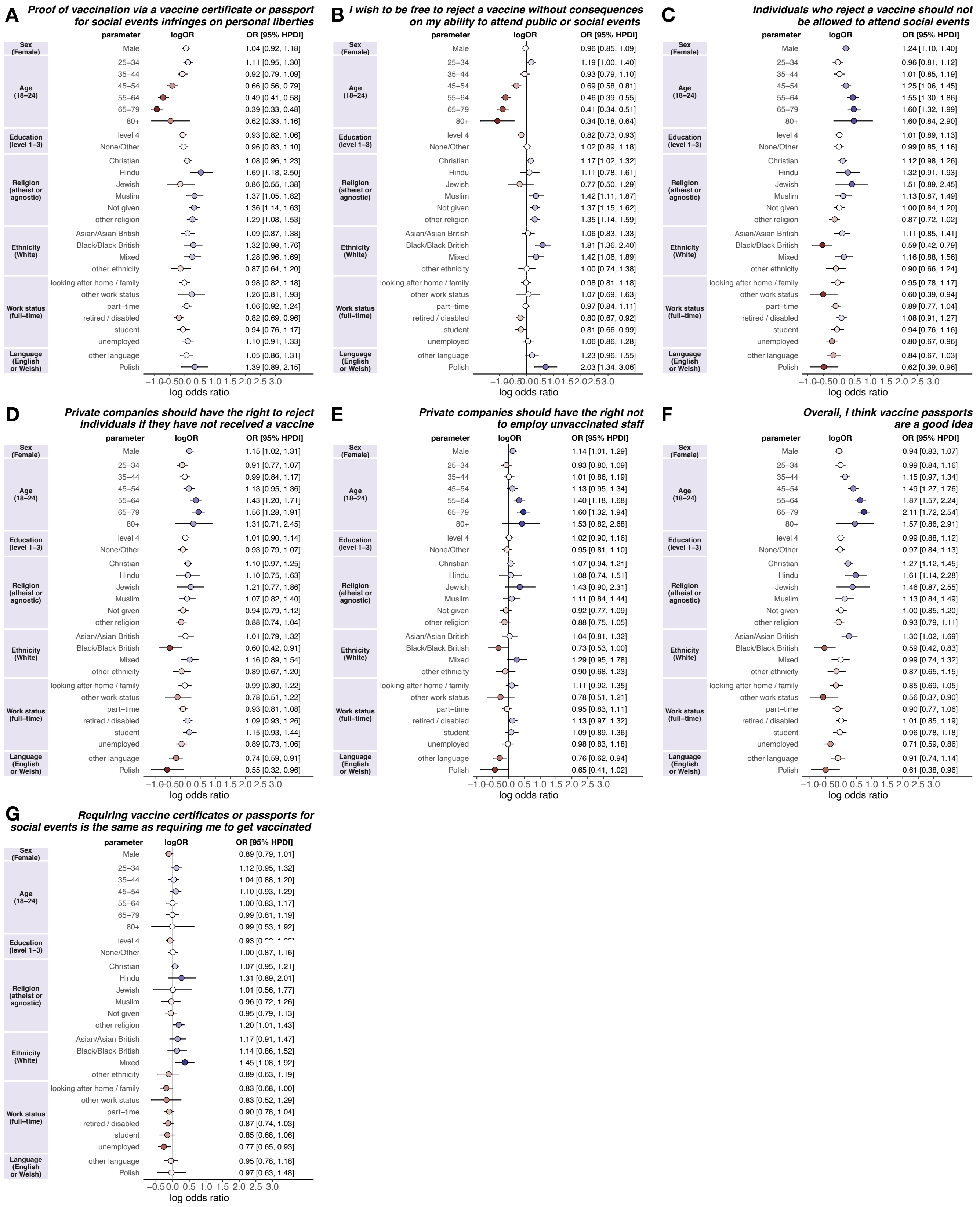


**Figure 3 | Socio-demographic determinants of the seven-item battery on attitudes to vaccinations, passports, and societal freedoms** Multilevel regression fixed-effect parameter log odds ratios are plotted with corresponding 95% highest posterior density intervals. Log odds ratios are coloured by effect magnitude and direction, where blues (reds) signify that the group is more (less) inclined than the baseline group to accept a COVID-19 vaccine and the darker the colour the stronger the association. For each factor, the baseline group is provided in parentheses on the left. Odds ratios with 95% HPDIs are shown on the right for each parameter.

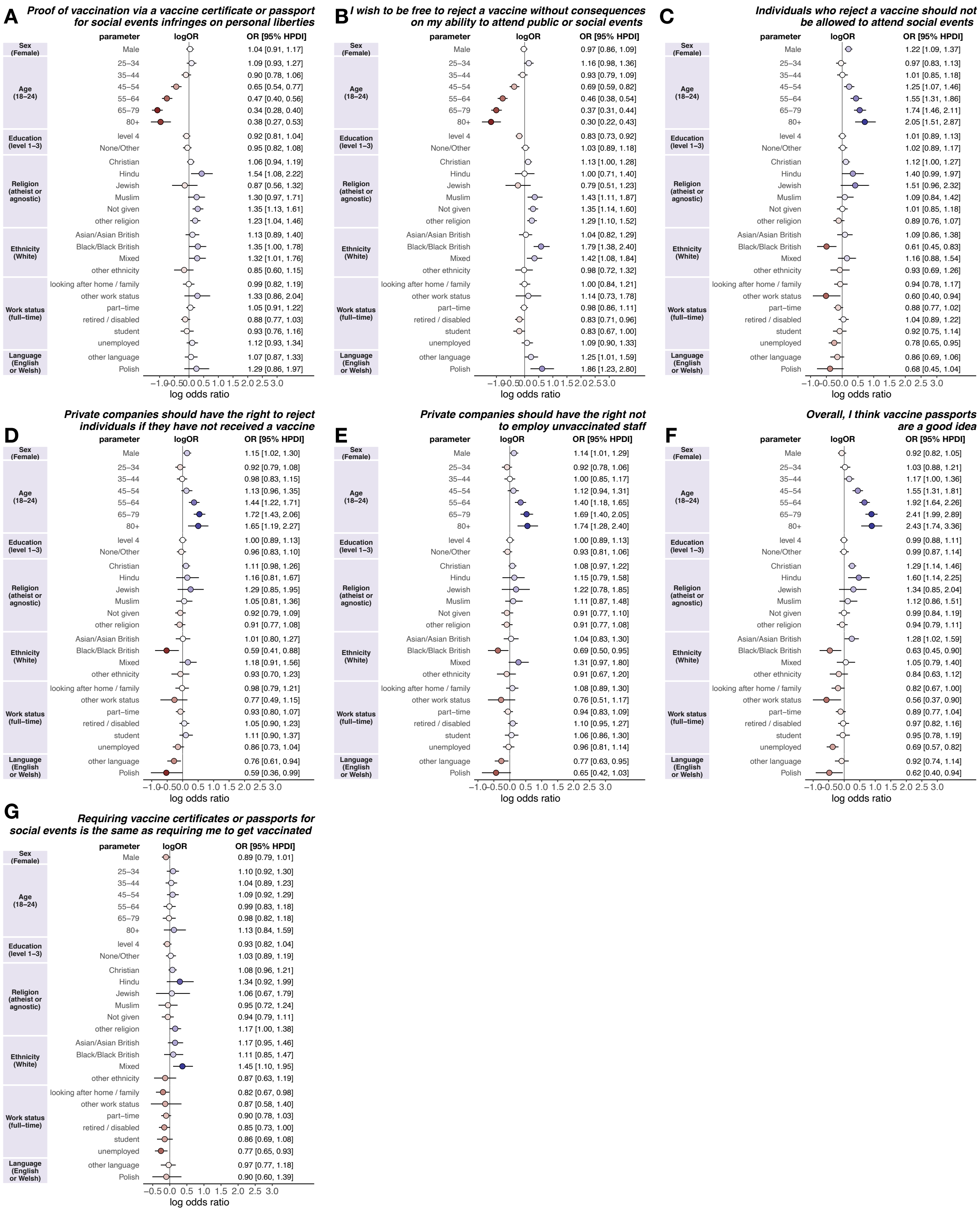

**Figure 4 | Socio-demographic determinants of the seven-item battery on attitudes to vaccinations, passports, and societal freedoms (sensitivity analysis)** Figures are as described in appendix, figure 3. All individuals are included in these regressions, but individuals who were previously removed for stating that they ‘prefer not to say’ (appendix, figure 3) are instead recoded to ‘neither agree nor disagree’. The results are quantitatively similar.

| | | | If a coronavirus (COVID-19) vaccine was required to attend social events in the UK (such as sports events, theatres, pubs, or restaurants), would you be more or less inclined to accept a coronavirus (COVID-19) vaccine? | | | | | | | | | | If a coronavirus (COVID-19) vaccine was required for international travel, would you be more or less inclined to accept a coronavirus (COVID-19) vaccine? | | | | | | | | | |
|---|---|---|---|---|---|---|---|---|---|---|---|---|---|---|---|---|---|---|---|---|---|---|
| | | | Much less likely | | Somewhat less likely | | Neither more nor less likely | | Somewhat more likely | | Much more likely | | Much less likely | | Somewhat less likely | | Neither more nor less likely | | Somewhat more likely | | Much more likely | |
| | | | n | % | n | % | n | % | n | % | n | % | n | % | n | % | n | % | n | % | n | % |
| Baseline vaccination intent | No, definitely not | 632 | 312 | 49.4 | 27 | 4.3 | 249 | 39.4 | 28 | 4.4 | 16 | 2.5 | 282 | 44.6 | 31 | 4.9 | 257 | 40.7 | 34 | 5.4 | 28 | 4.4 |
| | Unsure but lean no | 741 | 171 | 23.1 | 81 | 10.9 | 369 | 49.8 | 101 | 13.6 | 19 | 2.6 | 137 | 18.5 | 79 | 10.7 | 344 | 46.4 | 124 | 16.7 | 57 | 7.7 |
| | Unsure, but lean yes | 1717 | 134 | 7.8 | 170 | 9.9 | 786 | 45.8 | 431 | 25.1 | 196 | 11.4 | 110 | 6.4 | 143 | 8.3 | 727 | 42.3 | 429 | 25 | 308 | 17.9 |
| | Yes, definitely | 11453 | 267 | 2.3 | 290 | 2.5 | 5507 | 48.1 | 1645 | 14.4 | 3744 | 32.7 | 245 | 2.1 | 293 | 2.6 | 4875 | 42.6 | 1619 | 14.1 | 4421 | 38.6 |
| Sex | Female | 7555 | 449 | 5.9 | 273 | 3.6 | 3615 | 47.9 | 1172 | 15.5 | 2046 | 27.1 | 397 | 5.2 | 273 | 3.6 | 3195 | 42.3 | 1184 | 15.7 | 2506 | 33.2 |
| | Male | 6988 | 435 | 6.2 | 295 | 4.2 | 3296 | 47.2 | 1033 | 14.8 | 1929 | 27.6 | 377 | 5.4 | 273 | 3.9 | 3008 | 43 | 1022 | 14.6 | 2308 | 33 |
| Age | 18-24 | 1797 | 161 | 9 | 153 | 8.5 | 647 | 36 | 369 | 20.5 | 467 | 26 | 145 | 8.1 | 161 | 9 | 601 | 33.4 | 346 | 19.2 | 544 | 30.3 |
| | 25-34 | 2579 | 204 | 7.9 | 184 | 7.1 | 1109 | 43 | 464 | 18 | 618 | 24 | 178 | 6.9 | 170 | 6.6 | 996 | 38.6 | 475 | 18.4 | 760 | 29.5 |
| | 35-44 | 2686 | 195 | 7.3 | 94 | 3.5 | 1328 | 49.4 | 478 | 17.8 | 591 | 22 | 162 | 6 | 90 | 3.4 | 1192 | 44.4 | 477 | 17.8 | 765 | 28.5 |
| | 45-54 | 2687 | 156 | 5.8 | 71 | 2.6 | 1312 | 48.8 | 395 | 14.7 | 753 | 28 | 143 | 5.3 | 66 | 2.5 | 1207 | 44.9 | 346 | 12.9 | 925 | 34.4 |
| | 55-64 | 2314 | 98 | 4.2 | 44 | 1.9 | 1186 | 51.2 | 244 | 10.5 | 742 | 32.1 | 76 | 3.3 | 29 | 1.2 | 1055 | 45.6 | 284 | 12.3 | 870 | 37.6 |
| | 65-79 | 2445 | 70 | 2.9 | 21 | 0.9 | 1310 | 53.6 | 251 | 10.3 | 793 | 32.4 | 70 | 2.9 | 28 | 1.1 | 1137 | 46.5 | 270 | 11 | 940 | 38.5 |
| | 80+ | 35 | 0 | 0 | 1 | 2.9 | 19 | 54.3 | 4 | 11.4 | 11 | 31.4 | 0 | 0 | 2 | 5.7 | 15 | 42.9 | 8 | 22.9 | 10 | 28.6 |
| Education | level 1-3 | 6461 | 401 | 6.2 | 288 | 4.5 | 2926 | 45.3 | 960 | 14.9 | 1886 | 29.2 | 374 | 5.8 | 264 | 4.1 | 2697 | 41.7 | 947 | 14.7 | 2179 | 33.7 |
| | level 4 | 6008 | 331 | 5.5 | 217 | 3.6 | 2999 | 49.9 | 969 | 16.1 | 1492 | 24.8 | 260 | 4.3 | 215 | 3.6 | 2619 | 43.6 | 960 | 16 | 1954 | 32.5 |
| | none/other | 2074 | 152 | 7.3 | 63 | 3 | 986 | 47.5 | 276 | 13.3 | 597 | 28.8 | 140 | 6.8 | 67 | 3.2 | 887 | 42.8 | 299 | 14.4 | 681 | 32.8 |
| Religious affiliation | Atheist or agnostic | 4331 | 211 | 4.9 | 146 | 3.4 | 2292 | 52.9 | 635 | 14.7 | 1047 | 24.2 | 179 | 4.1 | 107 | 2.5 | 2134 | 49.3 | 609 | 14.1 | 1302 | 30.1 |
| | Christian | 7097 | 414 | 5.8 | 245 | 3.5 | 3204 | 45.1 | 1060 | 14.9 | 2174 | 30.6 | 366 | 5.2 | 252 | 3.5 | 2803 | 39.5 | 1052 | 14.8 | 2624 | 37 |
| | Hindu | 195 | 17 | 8.7 | 16 | 8.2 | 54 | 27.7 | 48 | 24.6 | 60 | 30.8 | 13 | 6.7 | 18 | 9.2 | 54 | 27.7 | 41 | 21 | 69 | 35.4 |
| | Jewish | 88 | 5 | 5.7 | 2 | 2.3 | 36 | 40.9 | 7 | 8 | 38 | 43.2 | 6 | 6.8 | 7 | 8 | 27 | 30.7 | 8 | 9.1 | 40 | 45.5 |
| | Muslim | 611 | 44 | 7.2 | 53 | 8.7 | 240 | 39.3 | 133 | 21.8 | 141 | 23.1 | 38 | 6.2 | 68 | 11.1 | 189 | 30.9 | 137 | 22.4 | 179 | 29.3 |
| | not given | 954 | 75 | 7.9 | 46 | 4.8 | 478 | 50.1 | 144 | 15.1 | 211 | 22.1 | 68 | 7.1 | 46 | 4.8 | 438 | 45.9 | 158 | 16.6 | 244 | 25.6 |
| | other religion | 1267 | 118 | 9.3 | 60 | 4.7 | 607 | 47.9 | 178 | 14.1 | 304 | 24 | 104 | 8.2 | 48 | 3.8 | 558 | 44 | 201 | 15.9 | 356 | 28.1 |
| Ethnicity | Asian/Asian British | 936 | 58 | 6.2 | 73 | 7.8 | 366 | 39.1 | 178 | 19 | 261 | 27.9 | 54 | 5.8 | 73 | 7.8 | 289 | 30.9 | 188 | 20.1 | 332 | 35.5 |
| | Black/Black British | 394 | 58 | 14.7 | 37 | 9.4 | 146 | 37.1 | 79 | 20 | 74 | 18.8 | 49 | 12.4 | 36 | 9.1 | 119 | 30.2 | 74 | 18.8 | 116 | 29.4 |
| | Mixed | 301 | 26 | 8.6 | 31 | 10.3 | 107 | 35.5 | 56 | 18.6 | 81 | 26.9 | 24 | 8 | 28 | 9.3 | 101 | 33.5 | 63 | 20.9 | 85 | 28.2 |
| | White | 12645 | 716 | 5.7 | 415 | 3.3 | 6160 | 48.7 | 1846 | 14.6 | 3508 | 27.7 | 619 | 4.9 | 396 | 3.1 | 5580 | 44.1 | 1837 | 14.5 | 4213 | 33.3 |
| | other ethnicity | 267 | 26 | 9.7 | 12 | 4.5 | 132 | 49.4 | 46 | 17.2 | 51 | 19.1 | 28 | 10.5 | 13 | 4.9 | 114 | 42.7 | 44 | 16.5 | 68 | 25.5 |
| Work status | full-time | 6380 | 436 | 6.8 | 315 | 4.9 | 2790 | 43.7 | 1084 | 17 | 1755 | 27.5 | 370 | 5.8 | 299 | 4.7 | 2491 | 39 | 1081 | 16.9 | 2139 | 33.5 |
| | looking after home/family | 886 | 36 | 4.1 | 27 | 3 | 501 | 56.5 | 114 | 12.9 | 208 | 23.5 | 43 | 4.8 | 26 | 2.9 | 458 | 51.7 | 117 | 13.2 | 242 | 27.3 |
| | part-time | 2532 | 172 | 6.8 | 116 | 4.6 | 1203 | 47.5 | 407 | 16.1 | 634 | 25 | 141 | 5.6 | 101 | 4 | 1084 | 42.8 | 396 | 15.6 | 810 | 32 |
| | retired / disabled | 3105 | 113 | 3.6 | 29 | 0.9 | 1676 | 54 | 316 | 10.2 | 971 | 31.3 | 104 | 3.4 | 36 | 1.2 | 1496 | 48.2 | 337 | 10.8 | 1132 | 36.5 |
| | student | 710 | 46 | 6.5 | 39 | 5.5 | 292 | 41.1 | 137 | 19.3 | 196 | 27.6 | 41 | 5.8 | 50 | 7 | 244 | 34.4 | 140 | 19.7 | 235 | 33.1 |
| | unemployed | 845 | 67 | 7.9 | 38 | 4.5 | 406 | 48 | 136 | 16.1 | 198 | 23.4 | 63 | 7.5 | 28 | 3.3 | 389 | 46 | 126 | 14.9 | 239 | 28.3 |
| | other work status | 85 | 14 | 16.5 | 4 | 4.7 | 43 | 50.6 | 11 | 12.9 | 13 | 15.3 | 12 | 14.1 | 6 | 7.1 | 41 | 48.2 | 9 | 10.6 | 17 | 20 |
| Language | English or Welsh | 13664 | 795 | 5.8 | 492 | 3.6 | 6542 | 47.9 | 2049 | 15 | 3786 | 27.7 | 705 | 5.2 | 472 | 3.5 | 5914 | 43.3 | 2042 | 14.9 | 4531 | 33.2 |
| | Polish | 93 | 14 | 15.1 | 13 | 14 | 39 | 41.9 | 16 | 17.2 | 11 | 11.8 | 11 | 11.8 | 8 | 8.6 | 40 | 43 | 17 | 18.3 | 17 | 18.3 |
| | other language | 786 | 75 | 9.5 | 63 | 8 | 330 | 42 | 140 | 17.8 | 178 | 22.6 | 58 | 7.4 | 66 | 8.4 | 249 | 31.7 | 147 | 18.7 | 266 | 33.8 |

**Table 1 | Cross-tabulation of change in inclination to accept a COVID-19 vaccine against baseline vaccination intent and socio-demographic status**

**Table 2 | Fixed-effect regression parameters from multilevel ordinal logistic regression model $Y|Z$ where $Y$ is change in vaccination inclination due to vaccine passports for domestic use.** Parameters associated with the effect of $Y$ on $Z$ are not shown, but all effects' 95% HPD exclude zero for all 12 UK regions.

| group | variable | region | odds ratio |
|---|---|---|---|
| | Level 4 (versus level 1-3) | North West | 0.84 [0.70, 1.00] |
| | Level 4 (versus level 1-3) | Yorkshire and the Humber | 0.82 [0.68, 0.99] |
| | Level 4 (versus level 1-3) | East Midlands | 0.74 [0.61, 0.91] |
| | Level 4 (versus level 1-3) | South East | 0.73 [0.62, 0.86] |
| **Education** | Level 4 (versus level 1-3) | Scotland | 0.82 [0.69, 0.99] |
| | Asian/Asian British (versus White) | West Midlands | 1.43 [1.03, 2.05] |
| **Ethnicity** | Asian/Asian British (versus White) | London | 1.31 [1.01, 1.71] |
| | Christian (versus atheist/agnostic) | Yorkshire and the Humber | 1.23 [1.03, 1.52] |
| | Christian (versus atheist/agnostic) | West Midlands | 1.42 [1.15, 1.70] |
| | Christian (versus atheist/agnostic) | East of England | 1.28 [1.07, 1.55] |
| | Jewish (versus atheist/agnostic) | London | 1.96 [1.13, 3.54] |
| | Christian (versus atheist/agnostic) | South East | 1.26 [1.07, 1.49] |
| | Christian (versus atheist/agnostic) | Wales | 1.36 [1.09, 1.71] |
| **Religion** | Christian (versus atheist/agnostic) | Northern Ireland | 1.34 [1.03, 1.78] |
| | Male (versus female) | East Midlands | 0.80 [0.66, 0.98] |
| | Male (versus female) | London | 0.82 [0.70, 0.96] |
| | Male (versus female) | South East | 0.79 [0.67, 0.91] |
| **Sex** | Male (versus female) | Scotland | 0.83 [0.70, 1.00] |
| | unemployed (versus full-time) | Yorkshire and the Humber | 0.68 [0.47, 0.99] |
| | part-time (versus full-time) | London | 0.79 [0.64, 0.96] |
| | student (versus full-time) | London | 1.55 [1.12, 2.16] |
| | retired / disabled (versus full-time) | South West | 0.72 [0.55, 0.94] |
| **Work status** | looking after home / family (versus full-time) | Scotland | 0.67 [0.48, 0.94] |

**Table 3 | Fixed-effect regression parameters from multilevel ordinal logistic regression model $Y|Z$ where $Y$ is change in vaccination inclination due to vaccine passports for international travel.**
Parameters associated with the effect of $Y$ on $Z$ are not shown, but all effects' 95% HPD exclude zero for all 12 UK regions.

| group | variable | region | odds ratio |
|---|---|---|---|
| | 55-64 (versus 18-24) | North West | 1.35 [1.01, 1.72] |
| | 65-79 (versus 18-24) | Yorkshire and the Humber | 1.40 [1.03, 1.86] |
| | 45-54 (versus 18-24) | East Midlands | 1.38 [1.04, 1.82] |
| | 65-79 (versus 18-24) | East Midlands | 1.36 [1.00, 1.87] |
| | 55-64 (versus 18-24) | South West | 1.41 [1.04, 1.85] |
| **Age** | 65-79 (versus 18-24) | South West | 1.36 [1.02, 1.86] |
| | Level 4 (versus level 1-3) | East Midlands | 0.80 [0.65, 0.99] |
| **Education** | Level 4 (versus level 1-3) | London | 1.22 [1.02, 1.45] |
| | Asian/Asian British (versus White) | North West | 1.47 [1.03, 2.08] |
| | Asian/Asian British (versus White) | Yorkshire and the Humber | 1.48 [1.02, 2.16] |
| | Asian/Asian British (versus White) | West Midlands | 1.57 [1.12, 2.18] |
| | Asian/Asian British (versus White) | East of England | 1.68 [1.14, 2.47] |
| | Asian/Asian British (versus White) | London | 1.40 [1.09, 1.79] |
| **Ethnicity** | Asian/Asian British (versus White) | Scotland | 1.62 [1.08, 2.52] |
| **Language** | Other (versus English / Welsh) | South East | 1.88 [1.22, 2.89] |
| | Christian (versus atheist/agnostic) | North East | 1.32 [1.04, 1.67] |
| | Christian (versus atheist/agnostic) | Yorkshire and the Humber | 1.22 [1.00, 1.46] |
| | Christian (versus atheist/agnostic) | West Midlands | 1.30 [1.06, 1.58] |
| | Christian (versus atheist/agnostic) | London | 1.18 [1.00, 1.40] |
| | Christian (versus atheist/agnostic) | South East | 1.29 [1.09, 1.52] |
| | Christian (versus atheist/agnostic) | South West | 1.28 [1.06, 1.55] |
| **Religion** | Christian (versus atheist/agnostic) | Northern Ireland | 1.32 [1.01, 1.74] |
| | Male (versus female) | West Midlands | 0.81 [0.68, 0.99] |
| | Male (versus female) | London | 0.85 [0.72, 0.99] |
| | Male (versus female) | South East | 0.84 [0.72, 1.00] |
| | Male (versus female) | Wales | 0.70 [0.56, 0.90] |
| **Sex** | Male (versus female) | Scotland | 0.78 [0.66, 0.93] |
| | looking after home / family (versus full-time) | North West | 0.72 [0.53, 0.98] |
| | student (versus full-time) | London | 1.44 [1.08, 1.99] |
| | looking after home / family (versus full-time) | South East | 0.72 [0.55, 0.97] |
| | looking after home / family (versus full-time) | South West | 0.68 [0.48, 0.97] |
| **Work status** | looking after home / family (versus full-time) | Scotland | 0.59 [0.42, 0.81] |

**Table 4 | First COVID-19 doses administered by age and sex** Data for the number of first-dose COVID-19 vaccinations administered provided by NHS England (https://coronavirus.data.gov.uk/details/vaccinations, accessed 30 April 2021) and 2019 ONS mid-year population estimates from https://www.ons.gov.uk/peoplepopulationandcommunity/populationandmigration/populationestimates (accessed 30 April 2021)

| | Doses administered (NHS England) | | Population estimate (ONS) | | Doses administered per population estimate (%) | |
|---|---|---|---|---|---|---|
| **Age** | **Male** | **Female** | **Male** | **Female** | **Male** | **Female** |
| **45-49** | 1,236,456 | 1,361,695 | 1,839,293 | 1,876,519 | 67.2 | 72.6 |
| **50-54** | 1,721,858 | 1,758,450 | 1,926,928 | 1,980,533 | 89.4 | 88.8 |
| **55-59** | 1,730,833 | 1,755,668 | 1,809,613 | 1,861,038 | 95.6 | 94.3 |
| **60-64** | 1,507,365 | 1,533,599 | 1,527,238 | 1,584,597 | 98.7 | 96.8 |
| **65-69** | 1,285,168 | 1,350,404 | 1,352,800 | 1,443,940 | 95.0 | 93.5 |
| **70-74** | 1,292,381 | 1,409,747 | 1,330,150 | 1,449,176 | 97.2 | 97.3 |
| **75-80** | 924,404 | 1,059,725 | 900,710 | 1,039,976 | 102.6 | 101.9 |
| **80+** | 1,104,100 | 1,593,389 | 1,152,541 | 1,684,423 | 95.8 | 94.6 |

### Questionnaire

DEM We will begin by asking you some questions about yourself

JS

DEMAGENUM How old are you?

▼ 18 (1) ... 100 (83)

DEMREG Which UK region do you live in?

▼ East Midlands (1) ... Other (for example, Jersey, Guernsey, Isle of Man) (13)

DEMSEX I am

- Male (1)
- Female (2)
- Other (3)

DEMEDU What is the highest level of education you have completed? (Select the response that best applies)

- No academic qualifications (1)
- 0-4 GCSE, O-levels, or equivalents (2)
- 5+ GCSE, O-levels, 1 A level, or equivalents (3)
- Apprenticeship (4)
- 2+ A levels or equivalents (5)
- Undergraduate or postgraduate degree, or other professional qualification (6)
- Other (e.g. vocational, foreign qualifications) (7)
- Do not know (8)
- Do not wish to answer (9)

DEMWRK Which of the following best describes your work status 6 months ago?

- ◯ Working full-time (including self-employed) (1)
- ◯ Working part-time (including self-employed) (2)
- ◯ Unemployed (3)
- ◯ Student (4)
- ◯ Looking after the home (5)
- ◯ Retired (6)
- ◯ Unable to work (including, for example, a short- or long-term disability) (9)
- ◯ Do not wish to answer (8)

---

DEMREL Do you consider yourself

- ◯ Christian (1)
- ◯ Hindu (2)
- ◯ Muslim (3)
- ◯ Jewish (4)
- ◯ Buddhist (5)
- ◯ Atheist or agnostic (6)
- ◯ Other (7)
- ◯ Do not wish to answer (8)

DEMETH Which best describes your ethnicity (select the response that best applies)

- White: English/Welsh/Scottish/Northern Irish/British (1)
- White: Irish (2)
- White: Other white background (3)
- White and Black Caribbean (4)
- White and Black African (5)
- White and Asian or White and Asian British (6)
- Black, African, Caribbean or Black British (12)
- Asian or Asian British: Indian (7)
- Asian or Asian British: Pakistani (8)
- Asian or Asian British: Chinese (10)
- Asian or Asian British: Other (11)
- Gypsy or Irish traveller (16)
- Other (13)
- Do not wish to answer (14)
- Roma (15)

---

DEMLAN What is your main language

- English or Welsh (1)
- Polish (2)
- Punjabi (3)
- Urdu (4)
- Bengali (5)
- Other (6)
- Do not wish to answer (7)

---

DEMINC What is your total household income in GBP (£) from all sources before tax?

- Under £15,000 (1)
- £15,000 to £24,999 (2)
- £25,000 to £34,999 (3)
- £35,000 to £44,999 (4)
- £45,000 to £54,999 (5)
- £55,000 to £64,999 (6)
- £65,000 to £99,999 (7)
- Over £100,000 (8)
- Do not wish to answer (9)

COV_INTRO **We will now ask you some questions about coronavirus (COVID-19) and new COVID-19 vaccines.**

---

COV_INV Have you received an invitation to receive a coronavirus (COVID-19) vaccine?

- ○ Yes (1)
- ○ No (2)
- ○ Do not know (3)

COV_DOSE Have you had at least one dose of a coronavirus (COVID-19) vaccine?

- ○ Yes, I have had one dose (1)
- ○ Yes, I have had both doses (2)
- ○ No (3)

COV_DOSE_2 Do you intend on receiving your second dose?

- ○ Yes, definitely (1)
- ○ Unsure, but leaning towards yes (2)
- ○ Unsure, but leaning towards no (3)
- ○ No, definitely not (4)

COV_VAC_1 Do you intend on accepting a coronavirus (COVID-19) vaccine?

- ○ Yes, definitely (1)
- ○ Unsure, but leaning towards yes (3)
- ○ Unsure, but leaning towards no (4)
- ○ No, definitely not (2)

COV_APT Have you booked an appointment to receive a coronavirus (COVID-19) vaccine?

- ○ Yes (1)
- ○ No (2)

COV_ACC_2 When you are invited to take a coronavirus (COVID-19) vaccine, will you accept the vaccine for yourself?

- ○ Yes, definitely (1)
- ○ Unsure, but leaning towards yes (2)
- ○ Unsure, but leaning towards no (3)
- ○ No, definitely not (4)

VAC_PASS

We would now like to ask you some questions about a vaccine or immunity certificate (commonly referred to as a "vaccine passport").

**A vaccine or immunity certificate is a physical or electronic document that confirms your status against a particular disease. For example, the certificate could confirm that you have been vaccinated against a disease or that you have some pre-existing immunity. Such a certificate could be saved on a mobile phone application.**
How strongly do you agree or disagree with the following statements? Social events refer to events such as sports events, theatres, pubs, or restaurants.

| | **Strongly disagree** (1) | **Tend to disagree** (2) | **Neither agree nor disagree** (8) | **Tend to agree** (3) | **Strongly agree** (4) | **Prefer not to say** (5) |
|---|---|---|---|---|---|---|
| Proof of vaccination via a vaccine certificate or passport for social events infringes on personal liberties (4) | ○ | ○ | ○ | ○ | ○ | ○ |
| I wish to be free to reject a vaccine without consequences on my ability to attend public or social events (5) | ○ | ○ | ○ | ○ | ○ | ○ |
| Individuals who reject a vaccine should not be allowed to attend social events (6) | ○ | ○ | ○ | ○ | ○ | ○ |
| Private companies should have the right to reject individuals if they have not received a vaccine (7) | ○ | ○ | ○ | ○ | ○ | ○ |
| Private companies should have the right not to employ unvaccinated staff (9) | ○ | ○ | ○ | ○ | ○ | ○ |
| Overall, I think vaccine passports are a good idea (12) | ○ | ○ | ○ | ○ | ○ | ○ |
| Requiring vaccine certificates or passports for social events is the same as requiring me to get vaccinated (13) | ○ | ○ | ○ | ○ | ○ | ○ |

VAC_PASS_UK If a coronavirus (COVID-19) certificate or passport was required to attend social events in the UK (such as sports events, theatres, pubs, or restaurants), would you be more or less inclined to accept a coronavirus (COVID-19) vaccine?

- ○ Much less inclined (1)
- ○ Somewhat less inclined (2)
- ○ Neither more nor less inclined (7)
- ○ Somewhat more inclined (8)
- ○ Much more inclined (5)
- ○ Do not know / prefer not to say (6)

---

VAC_PASS_INT If a coronavirus (COVID-19) certificate or passport was required for international travel, would you be more or less inclined to accept a coronavirus (COVID-19) vaccine?

- ○ Much less inclined (1)
- ○ Somewhat less inclined (2)
- ○ Neither more nor less inclined (3)
- ○ Somewhat more inclined (4)
- ○ Much more inclined (5)
- ○ Do not know / prefer not to say (6)
